# Tracing Early Molecular Drivers of Gastrinoma in MEN1: A Systematic Review for Precision Surveillance and Improved Patient Outcomes

**DOI:** 10.1101/2025.06.18.25329871

**Authors:** Ovais Shafi, Aakash, Luqman Naseer Virk, Mustafa Ali Hamid, Sohaib Khalid, Deepak Kumar, Raveena, Deepika Kumari Kataria, Muhammad Danial Yaqub, Rahimeen Rajpar, Manwar Madhwani, Faryal Yaqoob

**Affiliations:** Ovais Shafi, MBBS Sindh Medical College - Jinnah Sindh Medical University / Dow University of Health Sciences, Karachi, Pakistan; Aakash, MBBS/MD Florida State University Cape Coral Hospital, USA; Luqman Naseer Virk, MBBS Sindh Medical College - Jinnah Sindh Medical University / Dow University of Health Sciences, Karachi, Pakistan; Mustafa Ali Hamid, MBBS Lincoln County Hospital, Lincoln, United Kingdom; Sohaib Khalid, MBBS Lincoln County Hospital, Lincoln, United Kingdom; Deepak Kumar, MBBS/MD Zucker School of Medicine at Hofstra, Northwell at Mather Hospital, NY, USA; Raveena, MBBS Sindh Medical College - Jinnah Sindh Medical University / Dow University of Health Sciences, Karachi, Pakistan; Deepika Kumari Kataria, MBBS Chandka Medical College SMBBMU Larkana, Sindh, Pakistan; Muhammad Danial Yaqub, MBBS Lincoln County Hospital, Lincoln, United Kingdom; Rahimeen Rajpar, MBBS Sindh Medical College - Jinnah Sindh Medical University / Dow University of Health Sciences, Karachi, Pakistan; Manwar Madhwani, MBBS Sindh Medical College - Jinnah Sindh Medical University / Dow University of Health Sciences, Karachi, Pakistan; Faryal Yaqoob, PharmD Ziauddin University, Karachi Pakistan

**Author notes:** All authors have read and approved the manuscript. **Corresponding author: OS** Correspondence to Ovais Shafi.

**Keywords:** Gastrinomas, MEN1, Neuroendocrine Tumor, MEN1 Clinical Outcomes, Gastrinoma Risk Factors, High-risk MEN1 patients, Surveillance protocols, Biopsy-based detection, Risk stratification, Precision medicine, Therapeutic targeting

## Abstract

**Objective:** This study aims to investigate the increased risk in Multiple Endocrine Neoplasia type 1 patients to develop gastrinomas later in life. It focuses on the mechanisms that may come into play in duodenal enteroendocrine cells to result in gastrinoma development. By identifying key regulators involved, this study seeks to contribute towards future biomarker development and guide surveillance strategies in MEN1 patients to improve patient outcomes. Clinically, these insights may inform the development of gene expression–based surveillance tools that could be integrated into routine endoscopic or biopsy-based assessments for MEN1 patients.

**Background:** Gastrinomas, commonly associated with Multiple Endocrine Neoplasia type 1 (MEN1), are neuroendocrine tumors arising from duodenal enteroendocrine cells. The development and differentiation of these cells are also governed by transcription factors and signaling pathway. Loss of MEN1 disrupts this regulatory network, leading to cellular mis-specification, impaired differentiation, and increased risk of tumorigenesis. Understanding these early molecular events is clinically significant, as it may aid in identifying high-risk MEN1 patients, refining surveillance protocols, and guiding the development of targeted therapies for conditions such as Zollinger-Ellison syndrome.

**Methods:** Databases, including PubMed, MEDLINE, Google Scholar, and both open-access and subscription-based journals, were searched without date restrictions to investigate how the loss of MEN1 on developmental regulators (NEUROG3, ASCL1, PAX4, PAX6, ISL1, NKX2.2, INSM1, ARX, Notch, Wnt, BMP, Shh, MAPK/ERK, mTOR) of duodenal enteroendocrine cells, results in gastrinoma development. Studies meeting the criteria outlined in the methods section were systematically reviewed to address the research question. This study adheres to the PRISMA (Preferred Reporting Items for Systematic Reviews and Meta-Analyses) guidelines.

**Results:** Mutated MEN1 disrupts the expression of key transcription factors (NEUROG3, ASCL1, PAX4, PAX6, ISL1, NKX2.2, INSM1, ARX) and signaling pathways (Notch, Wnt, BMP, Shh, MAPK/ERK, mTOR) that govern the development and differentiation of duodenal neuroendocrine (enteroendocrine) cells. This dysregulation results in impaired cell fate specification, abnormal differentiation, and uncontrolled proliferation, events that collectively drive gastrinoma formation. These alterations may serve as early biomarkers for disease progression in MEN1 patients and offer potential targets for improved surveillance and personalized intervention strategies.

**Conclusion:** These findings highlight the critical role of MEN1 in maintaining epithelial homeostasis and suggest that molecular profiling of dysregulated transcription factors and signaling pathways may support early detection, risk stratification, and targeted surveillance in MEN1 patients. The loss of MEN1 impairs cell fate specification and differentiation, promoting abnormal proliferation and increasing the risk of gastrinoma formation. In the future, these insights may contribute to improved diagnostic strategies and the development of personalized therapeutic approaches, ultimately enhancing clinical outcomes for patients with MEN1-associated gastrinomas.

## Background

Gastrinomas are neuroendocrine tumors (NETs) that arise from enteroendocrine cells of the duodenum and are frequently associated with Multiple Endocrine Neoplasia Type 1 (MEN1), a hereditary tumor syndrome caused by mutations in the MEN1 tumor suppressor gene. Despite clinical advances in diagnosis and management, the molecular mechanisms by which MEN1 mutations drive duodenal gastrinoma formation remain incompletely understood [1]. Enteroendocrine cells originate from multipotent intestinal stem cells and their fate is directed by a tightly coordinated program involving a cascade of developmental transcription factors and signaling pathways, including NEUROG3, ASCL1, PAX4, PAX6, ISL1, NKX2.2, INSM1, and ARX, as well as modulatory input from pathways such as Notch, Wnt, BMP, Shh, MAPK/ERK, and mTOR. Each of these factors contributes to the proliferation, specification, and differentiation of neuroendocrine cells in the duodenum. However, how MEN1 loss interferes with this network remains poorly defined [2].

Understanding the impact of MEN1 inactivation on these developmental regulators is critical because MEN1-deficient duodenal neuroendocrine cells display a distinct phenotype characterized by altered differentiation and increased proliferative potential. It is increasingly recognized that tumorigenesis in MEN1 is not solely due to loss of cell cycle control but is also deeply rooted in lineage mis-specification and developmental arrest. By dissecting how MEN1 mutations disrupt key regulators of enteroendocrine development, this research provides essential insight into the early cellular events that drive gastrinoma formation [3]. This may help to identify the therapeutic vulnerabilities by targeting pathways that are hijacked during MEN1-deficient tumorigenesis. Therefore, this study is vital not only for the tumorigenesis landscape of gastrinoma but also for future strategies aimed at prevention, early detection, and guided treatment of duodenal neuroendocrine tumors [4].

This study significantly advances understanding of Zollinger-Ellison (ZE) syndrome by looking into the mechanisms involving the developmental regulators of enteroendocrine cells, that precede the formation of gastrinomas, which are the primary cause of ZE. By investigating how the loss of MEN1 disrupts a network of developmental regulators such as NEUROG3, ASCL1, PAX4, PAX6, ISL1, NKX2.2, INSM1, ARX, and key signaling pathways including Notch, Wnt, BMP, Shh, MAPK/ERK, and mTOR, the study focuses how abnormalities in enteroendocrine cell differentiation and fate specification contribute to hyperplastic and neoplastic processes within the duodenum [5]. This provides a developmental and transcriptional basis for the origin of gastrin-producing tumors. The insights from this study clarify how MEN1 loss shifts the balance from terminal differentiation toward unchecked proliferation of immature or mis-specified enteroendocrine progenitors, creating a cellular environment permissive for tumorigenesis [6, 7]. This developmental misregulation ultimately contributes to the hypergastrinemia that defines ZE syndrome. By linking MEN1 deficiency to early developmental disruptions rather than viewing gastrinoma formation as a late-stage proliferative event, the study redefines potential intervention windows. It opens opportunities for targeting upstream transcription factors and signaling regulators before overt tumor formation and symptomatic acid hypersecretion [8].

### Clinical Significance of this Study

Gastrinomas are among the most clinically challenging manifestations of Multiple Endocrine Neoplasia type 1 (MEN1), often leading to Zollinger-Ellison syndrome and severe gastrointestinal complications if not identified early. Despite regular surveillance in MEN1 patients, early detection of gastrinoma remains difficult due to its deep tissue origin, slow growth, and lack of reliable early biomarkers. This study addresses a critical gap by systematically analyzing the transcriptional and signaling dysregulation that results from MEN1 loss particularly the roles of NEUROG3, ASCL1, PAX4, and related pathways in disrupting enteroendocrine cell differentiation in the duodenum. By focusing on the early molecular events that precede overt tumor formation, this study highlights potential biomarkers for identifying high-risk individuals before clinical disease onset. Clinically, these insights may inform the development of gene expression–based surveillance tools that could be integrated into routine endoscopic or biopsy-based assessments for MEN1 patients. Furthermore, understanding the specific pathways altered by MEN1 loss may guide future research into chemopreventive or targeted therapies aimed at modulating differentiation and proliferation at the pre-neoplastic stage. Ultimately, this approach offers a more proactive model for MEN1 management, shifting from delayed detection and surgical intervention to timely risk stratification, early diagnosis, and potentially preventive treatment, thereby improving patient outcomes and long-term quality of life.

## Methods

### Aim of the Study

With focus on improving clinical outcomes for MEN Type1 patients at risk of gastrinoma development, this study investigates how the loss of MEN1 (menin) affects the transcriptional and signaling landscape of duodenal enteroendocrine progenitors, focusing on key regulators such as NEUROG3, ASCL1, PAX4, PAX6, ISL1, NKX2.2, INSM1, ARX, and associated signaling pathways including Notch, Wnt, BMP, Shh, MAPK/ERK, and mTOR. The goal is to look into the mechanisms by which MEN1 deletion alters enteroendocrine differentiation trajectories, resulting in neoplastic transformation and the development of gastrinomas.

### Research Questions

1. How does MEN1 loss affect the expression and function of developmental regulators involved in EEC lineage specification?
2. How does the disruption of these EEC developmental regulators contribute to gastrinoma formation and progression?

### Search Focus

A comprehensive literature search was conducted using the PUBMED database, MEDLINE database, and Google Scholar, as well as open access and subscription-based journals. There were no date restrictions for published articles. With focus on improving clinical outcomes for MEN Type1 patients at risk of gastrinoma development, the search strategy targeted:

- The role of MEN1 in regulating enteroendocrine progenitor gene networks
- EEC developmental regulators and their disruption in MEN1-deficient contexts
- The contribution of EEC developmental regulators dysregulation to gastrinoma tumorigenesis

Screening of the literature was also done on this same basis and related data was extracted. Literature search began in January 2021 and ended in December 2024. An in-depth investigation was conducted during this duration based on the parameters of the study as defined above. During revision, further literature was searched and referenced until April 2025. The literature search and all sections of the manuscript were checked multiple times during the months of revision (January 2025 – April 2025) to maintain the highest accuracy possible. This comprehensive approach ensured that the selected studies provided valuable insights into how mutated MEN1 impacts EEC developmental regulators and their impact on Gastrinoma tumorigenesis. This study adheres to relevant PRISMA guidelines (Preferred Reporting Items for Systematic Reviews and Meta-Analyses).

### Search Queries/Keywords

1. MEN1 and Enteroendocrine Development:

- “MEN1 and NEUROG3 in enteroendocrine cells”
- “ASCL1, PAX6, NKX2.2 expression in MEN1 knockout models”
- “MEN1 regulation of pancreatic and duodenal endocrine differentiation”
2. MEN1 and Developmental Signaling Pathways:

- “Notch and Wnt dysregulation in MEN1 loss”
- “BMP and Shh signaling in MEN1-deficient endocrine tissues”
- “MAPK/ERK and mTOR signaling in MEN1-related tumorigenesis”
3. MEN1 and Gastrinoma Mechanisms:

- “MEN1 mutations and gastrinoma formation”
- “Enteroendocrine dysregulation and neuroendocrine tumor development”
- “Gastrin-secreting tumors and transcriptional reprogramming in MEN1 syndrome”

Boolean operators (AND, OR) were used to refine search results. Additional searches were conducted to identify studies on how mutated MEN1 impacts EEC developmental regulators and their impact on gastrinoma tumorigenesis.

### Objectives of the Search

By identifying key regulators involved, this study seeks to contribute towards future biomarker development and guide surveillance strategies in MEN1 patients to improve patient outcomes. Clinically, these insights may inform the development of gene expression–based surveillance tools that could be integrated into routine endoscopic or biopsy-based assessments for MEN1 patients.

- To identify how MEN1 interacts with or represses key developmental regulators driving EEC development
- To track alterations in Notch, Wnt, BMP, and other developmental signals in MEN1-deficient tissues with focus on cell fate of EEC
- To understand how MEN1 loss predisposes enteroendocrine progenitors to tumorigenic reprogramming, particularly in the context of gastrinoma

### Screening and Eligibility Criteria

#### Initial Screening

Titles and abstracts were reviewed to ensure relevance to Mutated MEN1, Gastrinoma tumorigenesis, and EEC developmental regulators.

#### Full-Text Evaluation

Articles were further examined for detailed mechanistic insights into the involvement of genes, transcription factors and signaling pathways in Gastrinoma Development with focus on improving clinical outcomes for MEN Type1 patients at risk of gastrinoma development.

### Data Extraction Criteria

- The impact of MEN1 loss on the expression/function of EEC master regulators (e.g., NEUROG3, ASCL1)
- Pathway disruptions involving Notch, BMP, Wnt, MAPK, mTOR
- Direct links to gastrinoma development or EEC transformation

### Inclusion and Exclusion Criteria

#### Included

- Studies focused on MEN1 function in the EEC in duodenum
- Articles providing insights about EEC fate and its relation to MEN1
- Research linking MEN1 loss to Gastrinoma Tumorigenesis

#### Excluded

- Articles that did not conform to the study focus.
- Insufficient methodological rigor.
- Data not aligning with the research questions.

### Rationale for Inclusion

MEN1 acts as a tumor suppressor and transcriptional scaffold that regulates genes critical to neuroendocrine progenitor specification. Its loss results in transcriptional de-repression of mitogenic programs and dysregulation of progenitor TFs such as NEUROG3, ASCL1, and ISL1, while also interfering with Notch, Wnt, BMP, and Shh signaling, which are vital for cell fate restriction and terminal differentiation of EECs. Disruption of these programs contributes to lineage instability, promoting proliferation over differentiation and enhancing the risk of gastrinoma formation, particularly within the duodenal neuroendocrine compartment.

This comprehensive screening and inclusion rationale ensure that the selected studies provide valuable insights towards the objectives of the study. PRISMA Flow Diagram is Fig1.

### Assessment of Article Quality and Potential Biases

Ensuring the quality and minimizing potential biases of the selected articles were crucial aspects to guarantee the rigor and reliability of the research findings.

### Quality Assessment

The initial step in quality assessment involved evaluating the methodological rigor of the selected articles. This included a thorough examination of the study design, data collection methods, and analyses conducted. The significance of the study’s findings was weighed based on the quality of the evidence presented. Articles demonstrating sound methodology such as well-designed studies, controlled variables, and scientifically robust data were considered of higher quality. Methodological rigor served as a significant indicator of quality.

### Potential Biases Assessment

- **Publication Bias:** To address the potential for publication bias, a comprehensive search strategy was adopted to include a balanced representation of both positive and negative results, incorporating a wide range of published articles from databases like Google Scholar.
- **Selection Bias:** Predefined and transparent inclusion criteria were applied to minimize subjectivity in the selection process.

Articles were chosen based on their relevance to the study’s objectives, adhering strictly to these criteria. This approach reduced the risk of subjectivity and ensured that the selection process was objective and consistent.

- **Reporting Bias:** To mitigate reporting bias, articles were checked for inconsistencies or missing data. Multiple detailed reviews of the methodologies and results were conducted for all selected articles to identify and address any reporting bias.

By including high-quality studies and thoroughly assessing potential biases, this study aimed to provide a robust foundation for the results and conclusions presented.

### Language and Publication Restrictions

We restricted our selection to publications in the English language. There were no limitations imposed on the date of publication. Unpublished studies were not included in our analysis. By integrating insights from enteroendocrine developmental regulators and mutated MEN1, this study aims to advance the understanding of gastrinoma tumorigenesis.

## Results

A total of 3283 articles were identified using database searching, and 3122 were recorded after duplicates removal. 2781 were excluded after screening of title/abstract, 216 were finally excluded, and 5 articles were excluded during data extraction.

Finally, 120 articles were included as references.

### Clinical Relevance and Importance towards Improving Patient Outcomes

Gastrinomas in MEN1 patients remain a major clinical challenge due to their insidious onset, deep anatomical location, and frequent late detection often after significant progression or metastasis. Current surveillance protocols, while helpful, rely primarily on imaging and serum gastrin levels, which lack sensitivity for detecting early-stage or pre-neoplastic changes. This study provides a clinically significant advancement by identifying transcriptional and signaling dysregulation such as alterations in NEUROG3, ASCL1, PAX family genes, and Wnt/mTOR pathways that underlies the early stages of gastrinoma development in the duodenum following MEN1 loss. By systematically focusing on these early molecular disruptions, this research lays the groundwork for molecular-based surveillance tools that can detect high-risk changes before tumors become clinically apparent. The identification of transcriptional markers involved in cell fate mis-specification and unchecked proliferation offers potential biomarkers for stratifying MEN1 patients by risk level. This can enhance clinical decision-making by tailoring surveillance intensity, frequency, and therapeutic planning based on individual molecular profiles. Furthermore, these findings may inform the development of targeted therapies aimed at restoring normal cell differentiation or modulating affected pathways (e.g., Wnt, mTOR), potentially halting tumorigenesis at its earliest stages. Clinicians managing MEN1 patients particularly gastroenterologists, endocrinologists, and oncologists could ultimately integrate gene expression profiling into routine duodenal biopsies, transforming gastrinoma detection from a reactive process to a proactive, personalized approach. In doing so, this research directly supports the shift toward precision medicine, offering a path to earlier diagnosis, reduced tumor burden, and improved long-term outcomes in MEN1-associated gastrinomas.

### Investigating MEN1 Mutation-Induced Dysregulation of Duodenal Enteroendocrine Cells and Associated Gastrinoma Risk

## 1. NEUROG3

### Mutated MEN1 in Dysregulating Enteroendocrine Cells

NEUROG3 (Neurogenin 3) is a basic helix-loop-helix (bHLH) transcription factor that plays a pivotal role in the developmental biology of neuroendocrine and enteroendocrine cells in the gastrointestinal tract, particularly within the duodenum. During embryonic development, NEUROG3 is transiently expressed in progenitor cells and is indispensable for initiating the endocrine lineage specification in the gut epithelium. It activates a cascade of downstream transcription factors such as PAX4, NKX2.2, and NEUROD1, which drive the differentiation of multipotent progenitors into functionally distinct enteroendocrine cells capable of producing hormones like serotonin, cholecystokinin, and secretin [9]. The tightly regulated expression of NEUROG3 is critical for ensuring the proper timing and extent of endocrine cell commitment, and its expression is controlled by both extrinsic signals (such as Notch signaling, which represses its activity) and intrinsic epigenetic mechanisms that maintain the plasticity of intestinal stem cells. Loss of MEN1, a tumor suppressor gene encoding the nuclear scaffold protein menin, can lead to significant dysregulation of NEUROG3 expression, particularly in the context of duodenal neuroendocrine development and tumorigenesis. MEN1 is known to interact with various epigenetic regulators such as MLL (mixed-lineage leukemia) histone methyltransferases and HDACs, thus modulating chromatin accessibility and gene transcription. In the absence of functional MEN1, these regulatory interactions are disrupted, potentially leading to aberrant chromatin remodeling at the NEUROG3 promoter and enhancer regions [10]. This epigenetic dysregulation can result in either inappropriate reactivation of NEUROG3 in adult progenitor-like cells or sustained overexpression in committed endocrine cells, contributing to proliferative and neoplastic phenotypes. Moreover, menin has been implicated in the repression of bHLH transcription factors through modulation of H3K4 methylation status, suggesting that MEN1 loss may relieve this repression and lead to increased NEUROG3 transcription under non-physiological conditions. In addition, MEN1 loss may indirectly influence NEUROG3 through dysregulation of signaling pathways such as Wnt/β-catenin, Shh, or Notch, all of which have been implicated in intestinal homeostasis and neuroendocrine cell fate determination [11]. For example, enhanced Notch signaling maintains intestinal progenitors in an undifferentiated state by suppressing NEUROG3 expression, whereas suppression of Notch allows NEUROG3 induction and endocrine differentiation. If MEN1 deficiency alters Notch activity through downstream effectors like HES1, this could disturb the delicate balance of progenitor maintenance and endocrine commitment. Additionally, increased proliferation and survival of NEUROG3+ progenitor cells in MEN1-deficient contexts may reflect a broader failure to enforce terminal differentiation, allowing these cells to evade normal lineage constraints and potentially initiate tumorigenic programs. Overall, MEN1 loss creates a permissive chromatin and signaling environment that may both enhance NEUROG3 expression beyond its developmental window and enable NEUROG3+ cells to acquire proliferative, progenitor-like characteristics. This aberrant expression and maintenance of NEUROG3 could serve as an initiating event or a permissive condition for the development of duodenal neuroendocrine tumors, particularly those exhibiting features of enteroendocrine cell lineage commitment [12].

### Consequences for Enteroendocrine Cells and Associated Gastrionoma Risk

The loss of MEN1 profoundly disrupts the transcriptional and epigenetic landscape that governs neuroendocrine cell differentiation in the duodenum, particularly by altering the expression and activity of key developmental regulators such as NEUROG3. NEUROG3 functions as a master regulator of enteroendocrine cell fate, directing pluripotent intestinal progenitor cells toward the neuroendocrine lineage [13]. Under normal conditions, NEUROG3 expression is transient and tightly regulated during early development, ensuring that only a specific subset of progenitor cells exit the cell cycle and commit to differentiation into specialized hormone-producing cells. However, in the absence of functional MEN1, this regulatory precision is lost, resulting in the aberrant maintenance or reactivation of NEUROG3 expression in cells that would otherwise follow a non-endocrine or terminally differentiated fate. MEN1 encodes menin, a tumor suppressor and chromatin-binding scaffold protein that coordinates with multiple epigenetic complexes to repress or fine-tune the expression of genes critical for proliferation and differentiation. Loss of MEN1 disrupts these regulatory interactions, likely leading to inappropriate chromatin accessibility and sustained NEUROG3 expression in the duodenal epithelium. This persistent or ectopic NEUROG3 activity can lock progenitor cells in a partially differentiated or hyperplastic state, favoring the generation of an expanded pool of endocrine-committed cells with abnormal proliferative capacity [14]. As NEUROG3 is sufficient to drive endocrine lineage initiation but not terminal differentiation on its own, its dysregulated expression in the MEN1-deficient context may produce endocrine precursor cells that escape normal cell cycle exit signals and continue to proliferate. This disruption in cell fate has direct implications for the development of duodenal gastrinomas, one of the hallmark tumors associated with MEN1 syndrome. Gastrinomas arise from G cells gastrin-secreting enteroendocrine cells normally derived from NEUROG3+ progenitors. In MEN1-deficient tissues, the inappropriate expansion of NEUROG3+ progenitor populations increases the probability of accumulating additional oncogenic hits within this lineage. Furthermore, menin loss not only disrupts the NEUROG3 regulatory program but may also synergize with upregulated gastrin signaling, Wnt/β-catenin activity, or inflammatory cues to promote tumorigenic transformation. The result is a duodenal microenvironment in which lineage-specifying signals are preserved, but the restraints on proliferation and terminal differentiation are lost, predisposing NEUROG3-expressing progenitors to neoplastic conversion. Moreover, the failure to silence NEUROG3 in MEN1-deficient adult tissues may lead to a recapitulation of fetal endocrine differentiation programs that are otherwise inappropriate in mature tissue [15]. This reactivation of developmental pathways creates a pool of partially differentiated, mitotically active cells that retain endocrine features but do not fully mature or cease dividing. Such a scenario is particularly conducive to tumorigenesis in the neuroendocrine compartment, especially in the duodenum where gastrin-producing cells are already under tight regulatory control by both local and systemic signals. Therefore, MEN1 loss promotes a permissive environment for gastrinoma development by dysregulating NEUROG3-dependent cell fate decisions, expanding the population of vulnerable endocrine progenitors, and removing key tumor suppressive checkpoints that ordinarily limit neuroendocrine cell proliferation [16].

### Possible Clinical Implications of NEUROG3 Dysregulation in MEN1 Patients: Toward Improved Diagnosis, Surveillance, and Therapeutics

The dysregulation of NEUROG3 in MEN1-deficient duodenal tissues represents a pivotal early molecular event that can be clinically leveraged to improve the management of patients at risk for gastrinoma development. As a master regulator of enteroendocrine cell fate, NEUROG3 plays a critical role in directing intestinal progenitor cells toward the neuroendocrine lineage, including gastrin-secreting G cells. Under physiological conditions, NEUROG3 expression is transient and tightly restricted to developmental windows. However, in MEN1-deficient contexts, this regulatory precision is lost, resulting in the aberrant reactivation or sustained overexpression of NEUROG3 in adult duodenal cells, which in turn leads to an expanded pool of proliferative, incompletely differentiated endocrine progenitors an ideal substrate for neoplastic transformation.

Clinically, this aberrant NEUROG3 activity offers a valuable molecular marker for early-stage detection of neoplastic potential in MEN1 patients. Integration of biopsy-based NEUROG3 expression profiling into endoscopic surveillance protocols could allow clinicians to detect dysregulated differentiation programs before overt tumor formation, providing a window of opportunity for early intervention. For example, duodenal biopsies obtained during routine upper endoscopy could be assessed for NEUROG3 mRNA or protein expression using qPCR or immunohistochemistry, allowing for risk stratification based on molecular activity rather than gross tumor presence. Patients exhibiting abnormal NEUROG3 expression could be flagged for intensified surveillance, earlier follow-up intervals, or potential enrollment in preventive therapeutic trials. Furthermore, the mechanistic role of NEUROG3 in sustaining proliferative, lineage-committed precursor cells suggests that therapeutic targeting of the NEUROG3 regulatory axis, including upstream epigenetic modulators (e.g., HDACs, MLL complexes) or interacting pathways like Notch and Wnt may offer novel avenues for chemoprevention in high-risk individuals. Agents that restore epigenetic repression of NEUROG3, or modulate Notch signaling to enforce terminal differentiation, could potentially limit the expansion of tumor-prone progenitor populations. In this way, NEUROG3 is not only a biomarker but also a targetable node in the disease pathway, bridging molecular insight and therapeutic innovation. Since NEUROG3-driven reactivation of fetal endocrine programs appears central to MEN1-related tumorigenesis, its detection may also enable the development of non-invasive screening tools, such as circulating RNA signatures or exosomal markers derived from duodenal epithelium. These could complement biopsy findings and provide longitudinal monitoring tools for MEN1 patients.

NEUROG3 dysregulation serves as a clinically actionable molecular signature of early gastrinoma risk in MEN1 patients. Its integration into precision surveillance strategies, biopsy-based diagnostics, and potentially targeted therapies holds promise for improving patient outcomes by shifting the paradigm from delayed tumor detection to proactive, individualized risk management.

## 2. ASCL1

### Mutated MEN1 in Dysregulating Enteroendocrine Cells

ASCL1 is a transcription factor essential for the commitment and differentiation of neuroendocrine cells, including enteroendocrine cells in the duodenum. During development, ASCL1 operates downstream of lineage-specifying signals such as NEUROG3 and plays a critical role in promoting endocrine differentiation by activating a network of genes involved in secretory lineage fate, hormone biosynthesis, and neuronal-like features of neuroendocrine cells. ASCL1 functions as both a differentiation driver and a proliferation regulator, with its expression tightly controlled to ensure appropriate timing and spatial restriction of endocrine lineage commitment. In the duodenal epithelium, ASCL1 marks progenitor cells transitioning toward mature enteroendocrine identities and coordinates with factors such as NEUROD1 and INSM1 to consolidate the neuroendocrine gene program [17]. The loss of MEN1 profoundly affects the regulation of ASCL1, primarily by disrupting the chromatin landscape and transcriptional control mechanisms that maintain cell identity and lineage fidelity. MEN1 encodes menin, a nuclear scaffold protein that interacts with histone-modifying enzymes such as MLL (for H3K4 methylation) and HDACs, thereby shaping gene expression through epigenetic modulation. In MEN1-deficient contexts, these chromatin-based regulatory mechanisms are impaired, leading to inappropriate activation or sustained expression of developmental transcription factors like ASCL1 [18]. The absence of menin-mediated repression can render the ASCL1 locus more transcriptionally permissive, even in differentiated or proliferative compartments where its expression would typically be downregulated or silenced. This epigenetic derepression may lead to the expansion of ASCL1-expressing endocrine precursors or the inappropriate reactivation of ASCL1 in progenitor cells, promoting a partial neuroendocrine program that is coupled with continued proliferation. Moreover, ASCL1 expression in the gut epithelium is normally modulated by Notch signaling, which inhibits endocrine differentiation by suppressing bHLH transcription factors such as NEUROG3 and ASCL1. MEN1 loss has been shown to alter Notch pathway dynamics, either through direct transcriptional effects or by influencing the expression of pathway regulators like HES1. If MEN1 deficiency results in diminished Notch activity, the repression of ASCL1 may be lifted prematurely, enhancing endocrine specification at the expense of balanced epithelial cell differentiation. This premature or excessive ASCL1 expression could lock progenitor cells in an intermediate endocrine state, driving aberrant proliferation and reducing their capacity for terminal differentiation. Furthermore, menin may also regulate ASCL1 indirectly through suppression of mitogenic pathways like Wnt or Shh, whose upregulation in MEN1-deficient cells could support the survival and expansion of ASCL1+ cells inappropriately [19]. Altogether, the loss of MEN1 undermines the strict temporal and spatial control of ASCL1 expression in the duodenum, potentially resulting in the persistence of a proliferative, neuroendocrine-committed progenitor population. This dysregulation contributes to an altered cell fate landscape in which neuroendocrine lineage markers are expressed in cells that should otherwise differentiate or exit the cell cycle. In this disrupted context, ASCL1 may act not only as a differentiation factor but also as an oncogenic driver by enabling survival and expansion of endocrine-committed cells that are predisposed to transformation. Such a scenario creates fertile ground for the development of neuroendocrine tumors, including gastrinomas, by maintaining a pool of ASCL1-expressing cells that are vulnerable to secondary oncogenic insults. Thus, MEN1 loss deregulates the ASCL1-driven transcriptional program, altering cell fate decisions and contributing to the initiation and progression of duodenal neuroendocrine neoplasms [20].

### Consequences for Enteroendocrine Cells and Associated Gastrionoma Risk

The loss of MEN1 significantly alters the regulatory architecture that governs ASCL1 expression, leading to profound consequences on the cell fate of neuroendocrine cells in the duodenum and contributing to an increased risk of gastrinoma development. ASCL1 is a central transcription factor that directs progenitor cells toward a neuroendocrine lineage by activating a cascade of genes essential for endocrine specification, hormone production, and secretory machinery. Under normal physiological conditions, ASCL1 expression is transient and finely tuned, ensuring that neuroendocrine progenitors properly exit the cell cycle and differentiate into mature enteroendocrine cells, such as gastrin-secreting G cells. However, in the absence of MEN1, this regulatory precision is disrupted, and ASCL1 expression can become dysregulated, both temporally and spatially [21]. MEN1 encodes menin, a nuclear scaffold protein that coordinates chromatin remodeling and transcriptional repression by interacting with histone methyltransferases and other epigenetic regulators. Menin plays a crucial role in maintaining the epigenetic silencing of developmental regulators like ASCL1 in contexts where their expression is no longer appropriate. When MEN1 is lost, the chromatin landscape surrounding the ASCL1 locus becomes permissive to aberrant transcription, leading to the persistent or ectopic expression of ASCL1 in progenitor and even proliferative cell populations. This misexpression of ASCL1 results in the expansion of an endocrine-committed progenitor pool that is resistant to terminal differentiation cues and retains proliferative potential, a key prerequisite for tumorigenesis [22]. ASCL1 not only initiates neuroendocrine fate but also sustains a progenitor-like state if not properly downregulated. In MEN1-deficient duodenal epithelium, this creates a situation where cells expressing ASCL1 continue to proliferate while partially executing the endocrine program. This hybrid state is particularly conducive to neoplastic transformation, as it maintains lineage identity while promoting expansion. Moreover, ASCL1 is known to interact with pro-growth pathways and can suppress cell cycle exit genes, further skewing the balance toward proliferation. The resulting pool of immature neuroendocrine progenitors becomes a fertile substrate for tumor formation, especially under the influence of additional mitogenic or inflammatory stimuli [23]. This aberrant regulation of ASCL1 is especially significant in the context of gastrinoma development, as gastrin-producing cells derive from ASCL1+ precursors. The sustained expression of ASCL1 in MEN1-deficient tissue can lead to the inappropriate expansion of G cell precursors or the reprogramming of nearby epithelial cells into a gastrin-expressing, endocrine-like phenotype. The loss of menin removes a key checkpoint that normally limits endocrine progenitor expansion and enforces terminal differentiation, thus enhancing the probability that ASCL1+ cells will acquire further oncogenic alterations. These changes create a permissive environment for gastrinoma initiation in the duodenum, where expanded, dysregulated endocrine precursors aberrantly produce gastrin and escape growth control mechanisms. Therefore, MEN1 loss-mediated dysregulation of ASCL1 disrupts the normal trajectory of neuroendocrine differentiation, allowing for the persistence and expansion of proliferative ASCL1+ progenitors. This disruption in cell fate fidelity contributes directly to the pathogenesis of gastrinomas by increasing the population of cells susceptible to transformation and by sustaining an immature endocrine phenotype prone to hypersecretion and growth. The interplay between epigenetic deregulation, altered ASCL1 expression, and disrupted cell fate commitment underpins a key mechanism through which MEN1 inactivation drives duodenal neuroendocrine tumorigenesis [24].

### Possible Clinical Implications of ASCL1 Dysregulation in MEN1 Patients: Advancing Diagnosis, Surveillance, and Therapeutics in Gastrinoma Risk

The dysregulation of ASCL1 in MEN1-deficient duodenal tissue represents a clinically significant mechanism contributing to the early stages of gastrinoma formation. ASCL1 is a critical transcription factor that mediates neuroendocrine cell fate decisions and supports endocrine differentiation from progenitor cells, including those giving rise to gastrin-secreting G cells. In healthy tissue, ASCL1 expression is tightly regulated, transient, and restricted to developmental windows. However, loss of MEN1 disrupts this control by altering the chromatin landscape, leading to the inappropriate reactivation or persistent expression of ASCL1 in adult epithelial and progenitor cell compartments. From a clinical perspective, this aberrant ASCL1 expression has direct implications for early detection and risk assessment in MEN1 patients. The presence of ASCL1+ proliferative endocrine-committed progenitors especially in the absence of terminal differentiation creates a molecular profile that precedes overt neoplastic transformation. Therefore, ASCL1 can serve as an early biomarker of gastric neuroendocrine dysregulation, particularly in duodenal biopsies obtained during routine endoscopic surveillance of MEN1 mutation carriers. Immunohistochemical staining or mRNA profiling for ASCL1 could be incorporated into a molecular biopsy panel, allowing clinicians to identify high-risk patients based on the presence of aberrant endocrine progenitor expansion even in histologically normal tissue. Furthermore, the role of ASCL1 in sustaining a partially differentiated, proliferative endocrine phenotype makes it an attractive target for precision surveillance and therapeutic intervention.

Therapeutic approaches aimed at modulating ASCL1 expression or restoring regulatory signals lost in MEN1-deficient contexts, such as Notch and epigenetic repressors could help enforce terminal differentiation and reduce the proliferative capacity of endocrine precursors. Such differentiation therapies may prevent the transition from ASCL1+ progenitor expansion to overt gastrinoma formation, especially in patients demonstrating persistent ASCL1 expression in the duodenum. Moreover, since ASCL1-positive precursors may contribute to both hypergastrinemia and eventual tumor development, monitoring ASCL1 status longitudinally could offer insight into disease progression and therapeutic response. This would allow for dynamic surveillance strategies, where molecular activity, not just tumor size or serum gastrin levels, guides clinical decision-making. ASCL1 becomes a key node in a biomarker-guided surveillance protocol tailored specifically to MEN1 patients.

The dysregulation of ASCL1 following MEN1 loss disrupts the normal differentiation trajectory of enteroendocrine cells, expands the population of proliferative neuroendocrine precursors, and creates a molecular landscape highly permissive to gastrinoma formation. Integrating ASCL1 assessment into biopsy-based diagnostics, molecular surveillance, and targeted therapeutic strategies offers a powerful pathway toward improving early detection, reducing tumor burden, and enhancing personalized care for MEN1 patients at risk of gastrinoma.

## 3. PAX4

### Mutated MEN1 in Dysregulating Enteroendocrine Cells

PAX4 is a paired box homeodomain transcription factor that plays a critical role in the specification and differentiation of neuroendocrine cell types within the gastrointestinal tract, particularly in the duodenum. During development, PAX4 functions downstream of lineage-initiating transcription factors such as NEUROG3 and works in a coordinated network with other transcriptional regulators like PAX6, NKX2.2, and ARX to influence the fate decisions of endocrine progenitors [25]. Specifically, PAX4 is known for its role in promoting the differentiation of certain subtypes of enteroendocrine cells by repressing alternate lineage fates and facilitating the expression of genes necessary for hormone biosynthesis and neuroendocrine function. In the context of the duodenal epithelium, PAX4 expression is tightly regulated in space and time to ensure the balanced generation of various enteroendocrine cell populations, including those producing insulin-like peptides, somatostatin, or gastrin. The loss of MEN1 disrupts this finely tuned regulatory system, potentially leading to the dysregulation of PAX4 expression. MEN1 encodes the tumor suppressor protein menin, which acts as a scaffold for chromatin remodeling complexes and transcriptional repressors, thereby playing a fundamental role in maintaining transcriptional fidelity and cell identity. Menin exerts its influence through interactions with histone methyltransferases such as MLL (mixed lineage leukemia), which modify chromatin at promoters and enhancers of target genes, either activating or repressing gene expression in a context-dependent manner. In the absence of menin, the epigenetic regulation of key developmental genes like PAX4 may be lost, leading to either inappropriate silencing or, conversely, unregulated activation of its transcription depending on the surrounding chromatin and cellular signals. In MEN1-deficient cells, the loss of menin-mediated repression could render the PAX4 promoter more susceptible to aberrant transcriptional activation by upstream factors, especially in progenitor or dedifferentiated cell populations. Alternatively, menin might be required for the proper maintenance of active chromatin marks at the PAX4 locus in contexts where its expression is needed, such that MEN1 loss results in PAX4 silencing in cells that would otherwise commit to endocrine differentiation [26]. This dual potential for dysregulation either ectopic expression in inappropriate cell types or loss of expression in committed progenitors can significantly perturb the balance of enteroendocrine lineage differentiation. Furthermore, MEN1 deficiency may disrupt the expression of upstream regulators of PAX4, such as NEUROG3 or NKX2.2, whose own transcription is dependent on chromatin accessibility and signaling pathways modulated by menin. The misregulation of PAX4 resulting from MEN1 loss has direct implications for the development and function of enteroendocrine cells. For example, aberrant PAX4 expression may favor the expansion of specific hormone-producing cell lineages while repressing others, thereby skewing the normal cellular composition of the duodenal epithelium. Additionally, sustained or ectopic expression of PAX4 in a proliferative environment can contribute to the persistence of endocrine progenitor-like cells with incomplete differentiation, a state associated with increased susceptibility to neoplastic transformation. This effect is particularly concerning given the ability of PAX4 to suppress alternate lineage commitment, potentially locking cells into an endocrine-biased, proliferative phenotype when combined with MEN1-associated chromatin deregulation. Therefore, MEN1 loss may disrupt the expression dynamics of PAX4 at multiple levels including epigenetic, transcriptional, and signaling, thereby compromising the normal developmental trajectory of duodenal neuroendocrine cells. The resulting imbalance in differentiation and proliferation contributes to a permissive environment for tumorigenesis, including the formation of hormone-secreting neuroendocrine tumors. Through this mechanism, the dysregulation of PAX4 serves as a potential mediator linking MEN1 deficiency to disrupted endocrine cell fate and increased tumor risk in the gastrointestinal tract [27, 28].

### Consequences for Enteroendocrine Cells and Associated Gastrionoma Risk

The loss of MEN1 profoundly affects the regulatory networks governing PAX4 expression, leading to disturbances in the specification and differentiation of neuroendocrine cells in the duodenum and potentially heightening the risk of gastrinoma development. PAX4 plays a pivotal role in driving endocrine progenitors toward specific neuroendocrine fates, functioning as a key determinant in the suppression of alternate lineages and in the activation of genes associated with hormone-producing enteroendocrine cells. In a normal developmental context, PAX4 expression is transient and highly coordinated with other transcriptional regulators such as NEUROG3 and NKX2.2, ensuring that progenitor cells undergo timely cell cycle exit and terminal differentiation into mature hormone-expressing cells.

However, in the absence of MEN1, this regulatory precision is disrupted, resulting in misregulated PAX4 expression patterns that skew lineage fate decisions and destabilize endocrine identity [29]. MEN1 encodes menin, a chromatin-binding protein and tumor suppressor that is essential for maintaining epigenetic stability at loci governing developmental gene expression. Menin interacts with various histone-modifying enzymes and transcriptional complexes to enforce lineage-specific gene programs and suppress inappropriate or oncogenic transcription. Without menin, the chromatin landscape surrounding the PAX4 gene becomes vulnerable to deregulated histone methylation patterns and abnormal transcription factor access, leading to either loss of expression where PAX4 is normally required or ectopic expression in progenitor cells that should be committed to non-endocrine fates. This breakdown in regulatory control impairs the fidelity of endocrine lineage specification and differentiation, allowing for the accumulation of undifferentiated or partially differentiated cells that retain proliferative potential. Aberrant PAX4 expression in MEN1-deficient duodenal cells may sustain a pool of endocrine progenitors that are locked in a developmentally immature state. These progenitors are primed for hormone production but fail to fully differentiate, a condition that not only undermines normal duodenal epithelial homeostasis but also establishes a pro-tumorigenic niche. Furthermore, PAX4 is known to repress the expression of alternative fate determinants such as ARX, and its sustained activation can suppress pathways critical for lineage balance, thereby expanding gastrin-producing precursors at the expense of other cell types. In the context of MEN1 loss, this imbalance can favor the hyperplasia and eventual transformation of gastrin-expressing G cell precursors, which are especially susceptible to tumorigenesis under conditions of unchecked proliferation and disrupted differentiation cues [30]. The contribution of misregulated PAX4 to gastrinoma risk is further compounded by the growth-promoting environment created by MEN1 deficiency. Menin normally restricts endocrine cell proliferation through transcriptional repression of mitogenic pathways and by supporting the stability of cell cycle inhibitors. Its loss removes these brakes, allowing PAX4-expressing progenitors to persist and expand abnormally. This persistence of PAX4-positive, gastrin-lineage-biased cells provides a substrate for additional oncogenic events, ultimately increasing the likelihood of gastrinoma formation. The combined effects of impaired cell fate resolution, expanded endocrine-biased progenitor pools, and pro-proliferative signaling in MEN1-deficient tissue highlight how PAX4 dysregulation acts as a central node in the cascade that links MEN1 loss to duodenal neuroendocrine tumorigenesis. MEN1 loss disrupts the precise epigenetic and transcriptional regulation required for PAX4-mediated endocrine cell differentiation, resulting in a breakdown of normal cell fate commitment within the duodenal epithelium. The sustained or ectopic expression of PAX4 in proliferative progenitor cells biases lineage outcomes toward gastrin-producing fates, while also preventing terminal differentiation. This shift in cellular dynamics, together with the tumor-permissive environment fostered by menin deficiency, promotes the emergence of gastrin-secreting neuroendocrine tumors and establishes PAX4 as a mechanistic link between MEN1 mutation and gastrinoma development [31, 32].

### Possible Clinical Implications of PAX4 Dysregulation in MEN1 Patients: Advancing Precision Surveillance and Early Intervention in Gastrinoma

PAX4, a transcription factor critical for enteroendocrine lineage specification, emerges as a key clinical target in the context of MEN1-associated neuroendocrine tumorigenesis. In the healthy duodenal epithelium, PAX4 tightly regulates the differentiation of endocrine progenitors into hormone-producing cells, including gastrin-secreting G cells. However, in MEN1-deficient states, loss of menin disrupts the epigenetic and transcriptional control of PAX4, resulting in aberrant expression patterns that skew cell fate commitment, sustain proliferative progenitor populations, and bias lineage outcomes toward gastrinoma-prone phenotypes. This dysregulation presents important clinical opportunities for improving the care of MEN1 patients. First, aberrant PAX4 expression may serve as a molecular biomarker for early endocrine lineage imbalance in duodenal biopsies before the onset of overt tumor formation. PAX4 mRNA or protein detection via immunohistochemistry, qPCR, or chromatin accessibility assays in routine endoscopic tissue samples could support a biopsy-based screening protocol to identify high-risk individuals who exhibit endocrine progenitor expansion or differentiation blockade. Particularly in MEN1 mutation carriers, monitoring PAX4 could facilitate precision surveillance, allowing clinicians to escalate follow-up intervals or initiate early interventions in patients demonstrating molecular signatures of gastrinoma risk. Moreover, since PAX4 not only promotes endocrine commitment but also represses alternate fate programs and may contribute to persistent proliferative states when overexpressed, it represents a potential therapeutic target. Restoring regulatory control of PAX4 via epigenetic therapy (e.g., HDAC inhibitors or histone methylation modulators) or indirect pathway modulation (e.g., Notch, Wnt inhibitors) could reinstate normal differentiation trajectories and limit the expansion of undifferentiated endocrine precursors. This approach may be particularly useful for chemoprevention in patients with MEN1 mutations who have not yet developed clinically detectable tumors but display molecular abnormalities. Additionally, understanding the role of PAX4 in favoring gastrin-producing G cell hyperplasia over other enteroendocrine subtypes allows for more nuanced risk stratification.

Patients with duodenal tissues showing sustained or ectopic PAX4 expression may be predisposed not only to neuroendocrine tumorigenesis but also to functional tumors like gastrinomas, which carry unique clinical challenges such as Zollinger-Ellison syndrome. Thus, PAX4 expression profiling could help differentiate between low-risk and high-risk endocrine pathologies, informing both surveillance and therapeutic planning.

PAX4 dysregulation represents a clinically actionable molecular event in the MEN1-associated gastrinoma pathway. Its early detection in biopsies, potential as a therapeutic target, and role in defining endocrine progenitor dynamics make it a powerful biomarker for early diagnosis, personalized surveillance strategies, and targeted intervention in MEN1 patients. Incorporating PAX4 into the molecular monitoring landscape of MEN1 care could shift clinical practice from reactive tumor management to proactive, precision-based prevention and early treatment.

## 4. PAX6

### Mutated MEN1 in Dysregulating Enteroendocrine Cells

PAX6 is a critical transcription factor involved in the differentiation and functional specification of enteroendocrine cells in the duodenum, acting downstream of lineage commitment factors such as NEUROG3 and interacting with other regulators like PAX4, NKX2.2, and ARX. It plays a central role in defining terminal endocrine identity, promoting the transcription of genes necessary for hormone production and secretion, including those involved in gastrin, somatostatin, and serotonin expression. During duodenal development, PAX6 ensures the progression of committed endocrine progenitors toward mature hormone-secreting cell types and is essential for the transcriptional activation of key neuroendocrine markers [33]. The spatial and temporal regulation of PAX6 is tightly controlled by signaling pathways and chromatin states that define cellular competence and lineage outcome in the gut epithelium. The loss of MEN1 disrupts this precise regulation and can profoundly alter the expression dynamics of PAX6 in the duodenum. MEN1 encodes menin, a nuclear scaffold protein that interacts with histone methyltransferases (e.g., MLL complexes) and transcription factors to regulate gene expression through chromatin remodeling and epigenetic modifications.

Menin is crucial for maintaining transcriptional integrity at loci controlling differentiation and proliferation, including those involved in endocrine lineage progression. In the absence of menin, epigenetic control over the PAX6 locus may be compromised, leading to either a loss of activating histone marks needed for PAX6 expression or the inappropriate gain of repressive marks such as H3K27me3 mediated by polycomb repressive complexes. This could result in the silencing of PAX6 in endocrine progenitors that require its activity to complete their differentiation trajectory.

Moreover, menin indirectly affects PAX6 expression through its influence on upstream regulatory networks. MEN1 loss can perturb the expression of NEUROG3 and PAX4, both of which are known to regulate or interact with PAX6 either through direct transcriptional control or through modulation of cell identity states. For instance, if MEN1 loss leads to sustained or ectopic expression of PAX4, which is known to repress PAX6 under certain contexts, this could shift lineage decisions away from PAX6-dependent cell fates [34]. Additionally, MEN1 deficiency is associated with aberrant activation of growth-promoting pathways such as mTOR and Wnt signaling, which can disrupt the balance between progenitor maintenance and differentiation, further interfering with PAX6 transcriptional activation and stability. The dysregulation of PAX6 resulting from MEN1 loss undermines the fidelity of enteroendocrine lineage commitment and impairs the generation of fully differentiated, hormone-expressing cells. Without proper PAX6 expression, cells may remain in a progenitor-like state or differentiate aberrantly into nonfunctional or mis-specified endocrine populations. This failure to establish terminal identity not only affects hormonal homeostasis in the gut but also contributes to a proliferative milieu of endocrine-biased but developmentally stalled cells, setting the stage for tumor initiation and progression. PAX6’s role in maintaining differentiated neuroendocrine identity, its loss or misregulation due to MEN1 deficiency could allow for clonal expansion of immature cells, increasing susceptibility to transformation under additional genetic or environmental stressors. Thus, PAX6 represents a critical node through which MEN1 maintains normal endocrine differentiation, and its dysregulation serves as a plausible mechanistic link between MEN1 mutation and the development of neuroendocrine neoplasia in the duodenum [35, 36].

### Consequences for Enteroendocrine Cells and Associated Gastrionoma Risk

The loss of MEN1 has profound consequences on the transcriptional regulation of PAX6, a key transcription factor required for terminal differentiation and identity stabilization of neuroendocrine cells in the duodenum. PAX6 plays a critical role downstream of lineage-specifying factors such as NEUROG3, promoting the expression of genes necessary for the production and secretion of gut hormones including gastrin. In normal development, PAX6 ensures the maturation of enteroendocrine progenitors into specialized hormone-producing cells and restricts proliferation by reinforcing cell cycle exit and terminal fate commitment [37]. However, in MEN1-deficient conditions, this process is disrupted. Menin, the protein product of MEN1, is essential for maintaining transcriptional fidelity and chromatin stability at endocrine regulatory loci, including that of PAX6. Menin functions as a scaffold that facilitates histone methylation by MLL complexes and stabilizes the transcriptional activation of genes that promote differentiation. Its loss leads to widespread epigenetic deregulation, including aberrant histone modifications that may silence PAX6 expression or destabilize its transcriptional activity in endocrine progenitor cells. In the absence of menin, cells that would otherwise activate PAX6 and proceed toward a hormonally functional endocrine phenotype may instead retain a progenitor-like, partially specified state. This arrest in differentiation promotes the accumulation of immature endocrine cells that have not fully committed to a defined hormonal identity, including those biased toward gastrin-producing fates. The improper silencing or misexpression of PAX6 under such conditions impairs the transcriptional networks required for maturation and may simultaneously remove the suppressive effects that PAX6 exerts on proliferative signaling pathways, thereby creating a permissive environment for hyperplasia and transformation.

Furthermore, MEN1 loss may shift the balance of upstream regulators such as PAX4 and NEUROG3 in ways that antagonize or fail to activate PAX6 [38]. For example, overexpression of PAX4 in a MEN1-null context could suppress PAX6 expression, leading to a skewed differentiation trajectory that favors proliferative gastrin-expressing precursors over fully matured neuroendocrine cells. This dysregulated cell fate environment directly contributes to the pathogenesis of gastrinoma. Gastrinomas, which are characterized by excessive and unregulated gastrin secretion, often arise from cells that have failed to properly undergo terminal differentiation but continue to express gastrin under abnormal transcriptional control. The loss of PAX6, which is crucial for restraining this process through its role in endocrine maturation, removes an essential gatekeeper that normally prevents the inappropriate expansion of gastrin-producing populations. These populations, now unchecked in both differentiation and proliferation due to MEN1 loss, are susceptible to neoplastic transformation. In this context, the downregulation or misregulation of PAX6 contributes not only to the disruption of normal endocrine cell identity but also to the creation of a pre-neoplastic endocrine compartment vulnerable to additional oncogenic insults. Thus, the loss of MEN1 impairs PAX6-dependent differentiation programs in the duodenum, leading to an expansion of poorly differentiated, gastrin-expressing progenitors and thereby increasing the risk of gastrinoma development [39, 40].

### Possible Clinical Implications of PAX6 Dysregulation in MEN1 Patients: Toward Enhanced Diagnosis, Surveillance, and Targeted Therapies for Gastrinoma

The transcription factor PAX6 plays a pivotal role in the terminal differentiation and identity stabilization of enteroendocrine cells in the duodenum. It functions downstream of key lineage drivers like NEUROG3 and works in coordination with regulators such as PAX4 and ARX to promote the full maturation of hormone-producing cells, including gastrin-secreting G cells. In the setting of MEN1 deficiency, the epigenetic and transcriptional regulation of PAX6 is significantly impaired, resulting in a failure to activate its expression in endocrine progenitors. This disruption leads to the accumulation of poorly differentiated, proliferative neuroendocrine cells, which represent a critical intermediate state in the pathogenesis of gastrinoma. This understanding has direct clinical relevance for MEN1 patient management. Loss or downregulation of PAX6 can serve as an early molecular indicator of disrupted endocrine maturation in the duodenum, detectable in duodenal biopsy specimens before visible tumor formation. PAX6 expression profiling, via immunohistochemistry or RT-qPCR, could be incorporated into biopsy-based diagnostic panels to identify MEN1 patients harboring transcriptional signatures suggestive of a pre-neoplastic state. Detecting diminished PAX6 expression in high-risk patients may facilitate early intervention, intensification of surveillance frequency, or inclusion in preventive therapeutic trials. Moreover, PAX6 normally reinforces terminal differentiation and restrains proliferation. Its loss therefore marks a dual vulnerability both incomplete maturation and unchecked growth that can be targeted therapeutically.

Pharmacologic strategies aimed at restoring PAX6 expression or activating its downstream gene program could drive endocrine progenitors toward terminal differentiation, thereby reducing the pool of transformation-prone cells. Alternatively, indirect modulation of upstream regulators like NEUROG3, or inhibition of antagonists such as overexpressed PAX4, may help restore balance within the transcriptional network governing neuroendocrine lineage progression. In addition, PAX6 dysregulation skews cell fate toward hyperplastic, gastrin-producing states, a hallmark of gastrinoma development. This lineage bias can be identified at the molecular level and used to stratify patients by tumor subtype risk (e.g., G cell–dominant transformation) or progression potential. Such insights could refine personalized surveillance protocols, particularly in MEN1 carriers who present without clinical symptoms but show molecular signs of endocrine mis-specification.

PAX6 represents a clinically actionable node within the disrupted differentiation landscape of MEN1-associated gastrinoma. Its loss of expression identifies at-risk cellular states, and its restoration offers a rational therapeutic goal. By integrating PAX6 profiling into diagnostic and surveillance pathways, clinicians can move toward a precision medicine approach, enabling earlier detection, better-informed risk stratification, and intervention strategies that prevent or delay gastrinoma onset in MEN1 patients.

## 5. Islet1

### Mutated MEN1 in Dysregulating Enteroendocrine Cells

ISL1 (Islet-1) is a crucial transcription factor involved in the development and differentiation of neuroendocrine cells, including those in the duodenum. It plays an essential role in the early stages of neuroendocrine cell fate commitment by regulating genes that are responsible for endocrine hormone production, including those related to insulin, glucagon, and somatostatin. In the duodenum, ISL1 works in concert with other key transcription factors like NEUROG3 to promote the differentiation of enteroendocrine cells that produce gut hormones such as gastrin, serotonin, and somatostatin [41]. ISL1’s expression is tightly regulated during development to ensure proper endocrine cell specification and functional maturation. It also contributes to the fine-tuning of the neuroendocrine lineage by influencing both cell proliferation and differentiation, as well as the maintenance of endocrine cell identity through adulthood. The loss of MEN1 disrupts the regulation of ISL1 expression, as menin, the product of the MEN1 gene, is an important transcriptional regulator that modulates gene expression through chromatin remodeling and histone modifications. Menin interacts with various transcription factors and epigenetic regulators to ensure proper gene expression during cell differentiation and lineage specification. In the absence of MEN1, chromatin remodeling becomes deregulated, which can result in inappropriate silencing or activation of genes, including those regulating ISL1. Specifically, without menin, the balance of activating and repressive histone marks at the ISL1 locus could be disrupted. This could lead to either the loss of ISL1 expression, preventing the differentiation of enteroendocrine cells, or to its ectopic expression, which may drive premature or inappropriate differentiation of endocrine cells, skewing their fate decisions [42]. Moreover, the loss of MEN1 can also indirectly impact ISL1 expression through alterations in upstream signaling pathways. Menin is known to interact with various signaling cascades that control cell fate, such as the Wnt, Notch, and MAPK pathways. These pathways are involved in regulating the expression of genes necessary for neuroendocrine differentiation, and their disruption due to MEN1 loss could result in aberrant activation or suppression of ISL1. For example, if MEN1 loss leads to the activation of pathways that promote proliferation, such as the mTOR pathway, this could shift the balance from differentiation to expansion of progenitor populations, where ISL1 expression may be either repressed or misregulated. Additionally, MEN1 loss might affect the coordination of transcriptional networks, reducing the ability of ISL1 to work synergistically with other transcription factors like NEUROG3 and PAX6, which are essential for the orderly progression of endocrine differentiation in the duodenum. The loss of MEN1 leads to a dysregulation of ISL1 expression in neuroendocrine cells of the duodenum by disrupting both its direct transcriptional regulation and the broader signaling networks that control its activity [43]. This dysregulation impairs the proper differentiation of enteroendocrine cells and could lead to the accumulation of poorly differentiated cells. Furthermore, such disruption in ISL1 expression could have significant implications for the development of gastrointestinal neuroendocrine tumors, as the failure to properly regulate ISL1 could promote the expansion of progenitor populations and increase the likelihood of neoplastic transformation, particularly in the context of conditions like gastrinoma [44].

### Consequences for Enteroendocrine Cells and Associated Gastrionoma Risk

The loss of MEN1 profoundly disrupts the expression and function of ISL1, a key transcription factor essential for the differentiation and maintenance of neuroendocrine cells in the duodenum. ISL1 plays a crucial role in regulating the development of enteroendocrine cells, guiding their differentiation into hormone-producing cells that are necessary for gastrointestinal function. In a healthy developmental context, ISL1 works alongside other transcription factors like NEUROG3 and PAX6 to ensure the proper lineage specification and functional maturation of these cells. However, in the absence of MEN1, this finely tuned regulation is disturbed, which has profound effects on both the differentiation and the fate determination of neuroendocrine progenitors in the duodenum [45]. MEN1 loss leads to dysregulation of ISL1 expression through multiple mechanisms. Menin, the product of the MEN1 gene, is a key regulator of transcriptional complexes that control chromatin structure and gene expression. It helps mediate histone modifications and facilitates the recruitment of transcriptional machinery at specific loci. In the absence of MEN1, this regulatory function is lost, leading to an altered chromatin landscape at the ISL1 locus. As a result, ISL1 may either fail to be properly activated, which could impair the differentiation of neuroendocrine cells, or its expression may become aberrantly upregulated, disrupting the balance of neuroendocrine differentiation. This dysregulated expression of ISL1 can prevent proper maturation of enteroendocrine cells, leading to the persistence of progenitor-like cells that have not fully committed to a hormone-producing phenotype. In this context, MEN1 loss can cause an imbalance in the development of enteroendocrine cells, promoting either expansion of undifferentiated progenitors or abnormal differentiation into less mature, proliferative endocrine cells [46]. The loss of MEN1 also indirectly affects ISL1 expression through the disruption of critical signaling pathways that regulate endocrine cell differentiation. Menin is known to interact with signaling cascades like Notch, Wnt, and MAPK pathways, all of which are involved in regulating neuroendocrine cell fate decisions. The loss of MEN1 may result in the dysregulation of these pathways, which could further disturb the regulatory environment needed for proper ISL1 function. For instance, disruption of the Notch signaling pathway due to MEN1 loss could lead to a failure in maintaining the balance between progenitor cell proliferation and differentiation, potentially resulting in a pool of cells that are biased toward proliferative, undifferentiated states rather than proper endocrine differentiation. Additionally, aberrant Wnt signaling could enhance cell proliferation, creating a permissive environment for ISL1 misexpression and leading to abnormal neuroendocrine cell expansion. The impact of MEN1 loss on ISL1 expression and function has direct implications for the development of gastrinoma, a tumor characterized by excessive gastrin production [47]. Gastrinoma arises from enteroendocrine cells that have failed to properly differentiate and become hyperproliferative. If MEN1 loss leads to an overexpression or misregulation of ISL1, this could promote the abnormal expansion of gastrin-producing cells. ISL1 is involved in regulating the hormonal identity of enteroendocrine cells, and its misregulation can lead to a population of cells that exhibit both proliferative characteristics and an aberrant differentiation trajectory. In the case of MEN1 loss, this could result in the inappropriate accumulation of gastrin-producing cells, which may undergo neoplastic transformation, thus increasing the risk of gastrinoma formation. Furthermore, the disruption of ISL1’s role in maintaining proper endocrine differentiation could result in the generation of cells that continue to proliferate unchecked, a hallmark of tumorigenesis. Consequently, the loss of MEN1 contributes to the dysregulation of ISL1 and other neuroendocrine differentiation factors, thereby fostering a cellular environment conducive to the development of gastrinoma in the duodenum [48].

### Possible Clinical Implications of ISL1 Dysregulation in MEN1 Patients: Enhancing Diagnosis, Surveillance, and Precision Management of Gastrinoma

ISL1 is a core transcriptional regulator of neuroendocrine cell differentiation and hormonal identity in the gastrointestinal tract. Its tightly regulated expression ensures that enteroendocrine progenitor cells in the duodenum mature into fully functional, hormone-producing cells. In the context of MEN1 deficiency, loss of menin-mediated epigenetic control leads to the dysregulation of ISL1, resulting in an accumulation of immature, proliferative endocrine progenitors with an aberrant or stalled differentiation trajectory. This disruption plays a central role in the early pathogenesis of gastrinomas, and its clinical implications are significant. From a diagnostic standpoint, ISL1 represents a promising biomarker for early detection of neuroendocrine dysregulation in MEN1 patients. Aberrant ISL1 expression, whether through loss in differentiating cells or ectopic upregulation in progenitor compartments, can be detected in duodenal biopsy specimens through immunohistochemistry or transcriptomic profiling. The identification of ISL1-positive, partially differentiated neuroendocrine populations in histologically normal tissue may indicate a molecularly high-risk state before tumor development. Thus, integrating ISL1 assessment into routine endoscopic biopsy analysis offers a pathway to improve early-stage detection and risk stratification for gastrinoma. In terms of surveillance, MEN1 patients showing persistent ISL1 misexpression could be enrolled in enhanced monitoring protocols, including more frequent endoscopic evaluation, molecular re-biopsy, or imaging. Tracking ISL1 expression longitudinally may serve as a surrogate marker for progression from endocrine progenitor expansion to neoplastic transformation, allowing for dynamic risk adjustment in personalized surveillance plans. On the therapeutic front, ISL1 dysregulation highlights a key vulnerability in the endocrine differentiation program that can be targeted pharmacologically. Agents that restore proper chromatin regulation such as HDAC inhibitors or modulators of MLL histone methyltransferase activity may correct the transcriptional imbalance at the ISL1 locus.

Furthermore, indirect approaches such as Notch pathway modulation, already known to influence ISL1 expression and neuroendocrine differentiation, may help drive terminal maturation of endocrine precursors and prevent tumorigenic progression. ISL1 misregulation appears to bias progenitor cells toward gastrin-producing fates, contributing to the hyperplasia and expansion of G-cell lineages, a defining feature of gastrinomas. Therefore, monitoring ISL1 in combination with other markers like NEUROG3, PAX6, and gastrin could offer a more comprehensive molecular signature of gastrinoma risk. This multiplexed approach would further improve diagnostic specificity and support the development of biomarker-guided preventive strategies in MEN1 patients.

ISL1 is both a mechanistic driver and a clinically actionable biomarker in MEN1-associated gastrinoma development. Its dysregulation provides critical insight into early tumorigenic pathways and offers multiple points of clinical intervention from biopsy-based molecular diagnostics and personalized surveillance to epigenetic reprogramming therapies. Incorporating ISL1 into MEN1 management algorithms represents a significant advancement toward precision medicine in gastroenteropancreatic neuroendocrine tumors.

## 6. NKX2.2

### Mutated MEN1 in Dysregulating Enteroendocrine Cells

NKX2.2 is a crucial transcription factor involved in the development and differentiation of neuroendocrine cells, including those in the duodenum. It plays a vital role in specifying the identity of enteroendocrine cells by regulating the expression of genes that control the differentiation of precursor cells into specialized hormone-producing cells. NKX2.2’s role is particularly important in the proper differentiation of cells producing hormones like insulin, glucagon, and serotonin, which are essential for maintaining gut homeostasis. During development, NKX2.2 ensures the establishment of the neuroendocrine lineage by promoting the expression of genes required for both differentiation and functional maturation of these cells [49]. Its activity is tightly regulated to ensure proper cellular identity and the correct timing of endocrine cell differentiation in the gastrointestinal tract. The loss of MEN1 disrupts the regulation of NKX2.2 expression through both direct and indirect mechanisms. Menin, the protein product of the MEN1 gene, acts as a transcriptional regulator and interacts with a variety of transcription factors and chromatin-modifying complexes to modulate gene expression. One of menin’s roles is to ensure proper chromatin remodeling and histone modifications at the loci of critical transcription factors like NKX2.2. In the absence of MEN1, this regulatory function is impaired, leading to the disruption of the chromatin landscape at the NKX2.2 gene locus. This can result in either insufficient activation or inappropriate silencing of NKX2.2 expression. If NKX2.2 is not properly activated, it can hinder the differentiation of neuroendocrine cells, preventing them from acquiring their full endocrine potential. On the other hand, if NKX2.2 is misregulated or overexpressed, it could drive abnormal differentiation or cause the expansion of undifferentiated or poorly differentiated cells, contributing to the loss of cellular diversity within the enteroendocrine population [50]. Moreover, the loss of MEN1 also leads to the dysregulation of key signaling pathways that interact with NKX2.2 during cell fate determination. Menin has known interactions with signaling pathways such as the Notch, Wnt, and MAPK pathways, all of which are involved in regulating the balance between progenitor cell proliferation and differentiation. These pathways are essential for maintaining the appropriate differentiation trajectory of enteroendocrine progenitors, and their disruption due to MEN1 loss can result in aberrant NKX2.2 expression. For example, altered Notch signaling, often observed in the absence of MEN1, could lead to the failure of progenitor cells to differentiate properly, promoting an accumulation of undifferentiated cells. Such progenitor cells may either fail to express NKX2.2 at the appropriate time or overexpress it in a way that drives premature differentiation or a failure to acquire a mature endocrine phenotype. This misregulation of NKX2.2 could also impair the establishment of proper gut hormone-producing cells, leading to a dysfunction in gastrointestinal endocrine regulation. The dysregulation of NKX2.2 expression due to MEN1 loss is particularly concerning in the context of neuroendocrine tumorigenesis. NKX2.2 is essential for the appropriate differentiation of enteroendocrine cells, and its misexpression can result in the unchecked proliferation of neuroendocrine progenitor cells. This could contribute to the formation of a precancerous or tumorigenic cell population, increasing the risk of developing neuroendocrine tumors, such as gastrinomas. Gastrinomas are typically characterized by an overproduction of gastrin, which leads to excessive acid secretion and subsequent gastrointestinal pathology. If NKX2.2 fails to regulate the proper differentiation of cells producing gastrin, it could promote the expansion of gastrin-secreting cells inappropriately, contributing to the formation of gastrinomas in the duodenum [51]. Moreover, the dysregulation of NKX2.2 could lead to the emergence of a heterogeneous and dysfunctional endocrine cell population, further increasing the likelihood of neoplastic transformation. The loss of MEN1 leads to the disruption of NKX2.2 expression, which in turn impacts the differentiation and fate of enteroendocrine cells in the duodenum. This dysregulation can impair the development of specialized hormone-producing cells and contribute to the expansion of undifferentiated progenitor populations, creating a cellular environment conducive to tumorigenesis. The misexpression of NKX2.2, whether through insufficient activation or overexpression, plays a critical role in the pathogenesis of neuroendocrine tumors like gastrinomas, underscoring the importance of MEN1 in maintaining the delicate balance of endocrine cell differentiation in the duodenum [52].

### Consequences for Enteroendocrine Cells and Associated Gastrionoma Risk

The loss of MEN1 significantly disrupts the role of NKX2.2 in the development and differentiation of neuroendocrine cells in the duodenum, leading to aberrations in the cellular fate of enteroendocrine progenitors and increasing the risk of gastrinoma. NKX2.2 is a key transcription factor responsible for the differentiation of neuroendocrine cells, including those that produce essential gastrointestinal hormones such as insulin, glucagon, and gastrin. In the normal developmental process, NKX2.2 ensures that enteroendocrine cells acquire the proper functional identity and maturity. However, when MEN1 is lost, the regulatory function of menin, which is crucial for chromatin remodeling and transcriptional regulation, is impaired [53]. This disruption in menin function affects NKX2.2’s expression, as menin plays an important role in mediating the chromatin environment at the NKX2.2 locus. The loss of MEN1 can lead to either inadequate activation of NKX2.2, preventing the differentiation of neuroendocrine cells, or to its inappropriate upregulation, which can lead to abnormal differentiation or the expansion of undifferentiated cells. Furthermore, the absence of MEN1 also affects key signaling pathways such as Notch, Wnt, and MAPK, which play a critical role in regulating the balance between progenitor cell proliferation and differentiation. These pathways are vital for guiding the correct differentiation of progenitor cells into mature enteroendocrine cells, and their disruption due to MEN1 loss can exacerbate NKX2.2 misexpression. Specifically, altered Notch signaling, often associated with MEN1 loss, can prevent progenitor cells from differentiating properly, causing a failure to activate NKX2.2 at the appropriate stages of differentiation [54]. Alternatively, the absence of MEN1 may lead to an increase in NKX2.2 expression, which can push progenitor cells toward premature differentiation or result in the formation of a heterogeneous and functionally abnormal population of neuroendocrine cells. The misexpression or dysregulation of NKX2.2 is particularly concerning in the context of neuroendocrine tumors such as gastrinomas. Gastrinomas are typically derived from neuroendocrine cells that have failed to properly differentiate and instead undergo uncontrolled proliferation. When NKX2.2 expression is improperly regulated, either due to insufficient expression or overexpression, it can lead to the expansion of proliferative progenitor cells that are not fully differentiated. This increase in proliferative cells can promote the formation of gastrin-producing cells, as NKX2.2 is involved in regulating the identity of endocrine cell populations. If the balance between differentiation and proliferation is disrupted, these gastrin-secreting cells may accumulate and form tumors, increasing the risk of gastrinoma development in the duodenum [55]. The loss of MEN1, by altering NKX2.2’s expression, thus contributes to an environment that favors the expansion of undifferentiated progenitors and the formation of neoplastic growths like gastrinomas, where the regulation of gastrin production is lost, leading to excessive acid secretion and pathological conditions in the gastrointestinal tract. This loss of MEN1 disrupts the proper function of NKX2.2, which is essential for the proper differentiation and maturation of enteroendocrine cells in the duodenum. The altered expression of NKX2.2, whether through underactivation or overexpression, impairs the fate specification of neuroendocrine cells, creating an environment that fosters the development of neuroendocrine tumors, including gastrinomas. The interplay between NKX2.2 and MEN1 is crucial in maintaining the delicate balance of cell differentiation and proliferation, and the loss of MEN1 results in dysregulated NKX2.2 expression that ultimately contributes to gastrointestinal tumorigenesis [56].

### Possible Clinical Implications of NKX2.2 Dysregulation in MEN1 Patients: Toward Improved Risk Stratification, Surveillance, and Targeted Intervention for Gastrinoma

NKX2.2 is a key transcriptional regulator in the development and functional maturation of enteroendocrine cells in the gastrointestinal tract. In normal physiology, it orchestrates the differentiation of progenitor cells into hormone-producing neuroendocrine cells, including those responsible for secreting gastrin. However, in the context of MEN1 loss, this tightly regulated transcriptional program is disrupted, leading to the misexpression or insufficient activation of NKX2.2, which impairs endocrine lineage specification and increases susceptibility to gastrinoma formation. From a clinical standpoint, dysregulated NKX2.2 expression serves as a molecular marker of early endocrine lineage disruption, making it a promising candidate for biopsy-based molecular diagnostics. In MEN1 mutation carriers undergoing surveillance, the presence of NKX2.2 dysregulation in duodenal biopsy samples, even in the absence of overt histological abnormalities, could signal a pre-neoplastic state marked by abnormal progenitor expansion or misdifferentiation. Immunohistochemistry or transcript-level analysis of NKX2.2 expression could thus be incorporated into routine endoscopic screening protocols to stratify patients by molecular risk level and enable timely clinical intervention. Furthermore, NKX2.2 misregulation contributes to a failure of endocrine progenitors to complete their maturation program, leaving behind a population of partially differentiated cells with proliferative potential. This shift creates a permissive environment for tumor initiation, particularly in the G-cell lineage from which gastrinomas arise. Clinicians may use NKX2.2 profiling alongside other markers such as NEUROG3, PAX4, and gastrin to define a high-risk molecular signature in MEN1 patients, supporting precision surveillance strategies that go beyond conventional imaging and serum biomarkers. Therapeutically, NKX2.2’s role in neuroendocrine differentiation highlights new opportunities for molecular targeting. For instance, modulation of upstream regulators or epigenetic mechanisms that restore balanced NKX2.2 activity such as Notch signaling correction or chromatin remodeling agents could shift the balance from proliferative, mis-specified progenitors to properly differentiated endocrine cells. This differentiation-based approach may reduce the pool of neoplastic precursors, acting as a chemopreventive strategy in patients with molecular signs of early tumorigenesis. Importantly, NKX2.2 dysregulation also underscores the need to monitor the functional heterogeneity of endocrine cell populations in MEN1 patients. Misexpression can lead to the emergence of poorly defined neuroendocrine subtypes with uncontrolled gastrin production, contributing to pathologies like Zollinger-Ellison syndrome even before frank tumor formation. Identifying and tracking such molecular alterations could guide early treatment planning, including the timing of surgical interventions or systemic therapy trials.

The disruption of NKX2.2 by MEN1 loss reflects a central mechanism in the initiation of duodenal neuroendocrine neoplasms, particularly gastrinomas. Monitoring NKX2.2 expression as part of a biopsy-based surveillance and risk assessment toolkit could improve early detection, refine risk stratification, and guide the development of targeted therapies that restore differentiation and suppress tumor progression. Integrating NKX2.2 into the clinical workflow of MEN1 management represents a step toward precision medicine in neuroendocrine tumor prevention and care.

## 7. INSM1

### Mutated MEN1 in Dysregulating Enteroendocrine Cells

INSM1 (insulinoma-associated protein 1) is a key transcription factor that plays a critical role in the development and differentiation of neuroendocrine cells, including those in the duodenum. It is crucial for the proper specification of enteroendocrine progenitors and for maintaining the identity of mature neuroendocrine cells. INSM1 is involved in regulating several aspects of neuroendocrine differentiation, including the promotion of hormone-producing cell types, such as those involved in gastrin production in the duodenum. The loss of MEN1, a tumor suppressor gene, can have significant consequences on the regulation of INSM1 and disrupt its role in neuroendocrine cell development.

MEN1 encodes the protein menin, which functions as a crucial co-regulator of transcription factors, and its loss leads to disruptions in multiple signaling pathways that regulate cell fate decisions in the duodenum. Menin has a well-established role in regulating the chromatin state of various target genes, including INSM1 [57]. Under normal circumstances, menin helps recruit chromatin-modifying complexes to the INSM1 promoter, ensuring that its expression is tightly regulated during the differentiation of neuroendocrine cells. When MEN1 is lost, menin’s ability to regulate INSM1 transcription is impaired. This loss of menin function can lead to either a reduction in INSM1 expression, preventing proper differentiation of neuroendocrine cells, or aberrant upregulation of INSM1, which could cause an overproduction of neuroendocrine cells and an increased risk of tumor formation. Dysregulation of INSM1 expression due to MEN1 loss can also affect the balance between progenitor cell proliferation and differentiation, as INSM1 is involved in maintaining the differentiation of progenitor cells into mature neuroendocrine cells. In addition to the direct impact on INSM1 transcription, MEN1 loss also influences several signaling pathways, such as Notch, Wnt, and MAPK, that intersect with INSM1 regulation. Notch signaling, in particular, is critical in maintaining the balance between progenitor cell proliferation and differentiation in the duodenum. Disruption of Notch signaling due to MEN1 loss could lead to premature differentiation of enteroendocrine progenitors, which may not properly mature into the correct subtypes of neuroendocrine cells. This aberrant differentiation process, coupled with dysregulated INSM1 expression, could lead to a population of neuroendocrine cells that are more prone to proliferation and tumorigenesis [58]. The altered regulation of INSM1, due to MEN1 loss, also has potential consequences for the development of neuroendocrine tumors such as gastrinomas. INSM1 is involved in maintaining the differentiation and functional identity of enteroendocrine cells that produce hormones like gastrin. In the absence of MEN1, the misregulation of INSM1 may lead to a failure in properly committing progenitor cells to their correct differentiated state, thereby favoring the expansion of undifferentiated or partially differentiated cells. These cells, particularly those in the duodenum, may begin to proliferate uncontrollably and form gastrin-producing tumors, such as gastrinomas. The overexpression or inappropriate activation of INSM1 can push the enteroendocrine cells towards a proliferative phenotype, increasing the likelihood of tumor formation and the pathological consequences of excessive gastrin secretion. The loss of MEN1 disrupts the regulation of INSM1, a critical transcription factor involved in the differentiation of neuroendocrine cells in the duodenum. MEN1 loss affects INSM1 expression by impairing menin’s co-regulatory function on chromatin, leading to either insufficient or excessive expression of INSM1. This dysregulation can disrupt the differentiation process of enteroendocrine progenitors and result in an accumulation of abnormal neuroendocrine cells, increasing the risk of developing gastrinomas [59]. The interplay between MEN1, INSM1, and various signaling pathways underscores the complexity of the regulatory network that governs neuroendocrine cell fate and highlights the potential for tumorigenesis when this network is disrupted [60].

### Consequences for Enteroendocrine Cells and Associated Gastrionoma Risk

The loss of MEN1 (Multiple Endocrine Neoplasia type 1), a tumor suppressor gene, significantly impacts the regulation of transcription factors that are crucial for the proper development and differentiation of neuroendocrine (enteroendocrine) cells in the duodenum. INSM1 (insulinoma-associated protein 1) is one of the key transcription factors involved in the differentiation of these cells, and its expression is tightly controlled by various regulatory networks, including those governed by MEN1. MEN1 encodes menin, a protein that plays a pivotal role in chromatin remodeling and transcriptional regulation. Menin acts as a cofactor for several transcription factors, and its loss leads to a loss of proper transcriptional regulation of target genes, including INSM1. The dysregulation of INSM1 expression resulting from the loss of MEN1 can disrupt the normal development of enteroendocrine cells, leading to an increased risk of developing neuroendocrine tumors, such as gastrinomas, in the duodenum. In normal development, INSM1 is expressed in enteroendocrine progenitor cells and is critical for their differentiation into mature hormone-producing cells, such as those producing gastrin. Menin, in its normal function, helps maintain the proper levels of INSM1 by acting as a co-regulator and ensuring its expression occurs at the right time and in the correct cells. When MEN1 is lost, the loss of menin’s regulatory influence on INSM1 leads to altered gene expression [61]. This can manifest as either insufficient INSM1 expression, leading to a failure in proper differentiation of neuroendocrine cells, or overexpression of INSM1, which can push progenitor cells into a proliferative state rather than allowing them to fully differentiate. Both outcomes are problematic because they disturb the balance between proliferation and differentiation in the neuroendocrine compartment of the duodenum, resulting in a higher risk of abnormal cell accumulation and potential tumor formation. The disruption of INSM1 expression due to MEN1 loss can also influence the developmental balance between different types of enteroendocrine cells. INSM1 is essential for the differentiation of a variety of neuroendocrine cell types, including those that produce hormones like gastrin, a key factor in gastric acid secretion [62]. When INSM1 expression is dysregulated, enteroendocrine progenitor cells may either fail to differentiate into the correct cell types or develop abnormally into proliferative, undifferentiated cells. These undifferentiated cells may continue to proliferate and, under the right conditions, form gastrinomas, a type of neuroendocrine tumor that typically arises in the duodenum. Gastrinomas are characterized by excessive gastrin secretion, which leads to an overproduction of gastric acid, causing peptic ulcers and other related complications. The altered differentiation program due to MEN1 loss means that cells with the potential for tumorigenesis may not undergo the normal maturation process, increasing the likelihood of tumor formation in the duodenum. Furthermore, MEN1 loss can also lead to disruption in other signaling pathways that regulate neuroendocrine differentiation, such as Notch and Wnt signaling [63]. These pathways interact with INSM1 and regulate the balance between progenitor cell self-renewal and differentiation. When MEN1 is lost, these pathways may become dysregulated, further skewing the developmental trajectory of enteroendocrine cells and contributing to an increased risk of tumor formation. The inability to properly regulate INSM1 in the context of these dysregulated signaling pathways can lead to an imbalance in the cell fate decisions of enteroendocrine progenitors, favoring uncontrolled cell growth and the formation of gastrinomas. The loss of MEN1 disrupts the regulation of INSM1, a key transcription factor involved in the differentiation of neuroendocrine cells in the duodenum. This disruption alters the normal cell fate of enteroendocrine progenitors, leading to either insufficient or excessive INSM1 expression. Such dysregulation increases the risk of abnormal cell proliferation, preventing proper differentiation and pushing the cells toward a proliferative, undifferentiated state. This accumulation of undifferentiated cells, particularly in the context of altered signaling pathways, significantly increases the likelihood of gastrinoma development, a neuroendocrine tumor associated with excessive gastrin production. Therefore, the loss of MEN1 represents a critical event that may predispose individuals to the development of gastrinomas by disrupting the normal regulatory mechanisms that control INSM1 expression and enteroendocrine cell differentiation [64].

### Possible Clinical Implications of INSM1 Dysregulation in MEN1 Patients: Advancing Early Diagnosis, Precision Surveillance, and Therapeutic Innovation in Gastrinoma Care

INSM1 is a central regulator of neuroendocrine differentiation, responsible for the transition of enteroendocrine progenitors into hormone-producing cells, including gastrin-secreting G cells in the duodenum. Its function is tightly controlled through epigenetic and transcriptional mechanisms, including those orchestrated by menin, the tumor suppressor encoded by MEN1. In MEN1-deficient states, this control is compromised, leading to aberrant INSM1 expression, which disrupts normal endocrine cell fate decisions and sets the stage for gastrinoma development.

Clinically, the dysregulation of INSM1 offers a powerful early molecular signal for identifying patients at risk of tumorigenesis before structural changes are detectable. MEN1 mutation carriers can benefit from biopsy-based INSM1 profiling, which could be incorporated into routine duodenal surveillance via endoscopy. Overexpression or underexpression of INSM1, especially in histologically normal tissue, may indicate underlying endocrine misdifferentiation and warrant closer clinical follow-up or earlier intervention. Its assessment alongside other transcriptional markers (e.g., NEUROG3, ISL1, PAX4) may allow for a multi-gene risk stratification model, enhancing the specificity and sensitivity of surveillance in MEN1 patients. From a diagnostic perspective, INSM1 is already being validated as a pan-neuroendocrine marker in various tumor types. Its expression in duodenal biopsies could therefore serve a dual role both as a screening tool for early pre-neoplastic changes and as a confirmatory marker in histologically ambiguous lesions. Such integration into the diagnostic workflow would help clinicians distinguish between benign hyperplasia, pre-invasive lesions, and early neuroendocrine tumors, guiding appropriate clinical decision-making. Therapeutically, INSM1 dysregulation points toward a targetable disruption in the neuroendocrine differentiation pathway. Patients with persistent INSM1 overexpression may benefit from differentiation-restoring agents, such as epigenetic modulators (e.g., HDAC inhibitors, histone methyltransferase inhibitors) that aim to rebalance cell fate programs. Additionally, understanding how INSM1 interacts with disrupted signaling cascades (Notch, Wnt, MAPK) offers avenues for combination therapies that suppress proliferation while promoting proper cell maturation. Functionally, INSM1 dysregulation contributes to the expansion of immature endocrine-like cells, many of which may retain the ability to express gastrin, but without undergoing normal terminal differentiation. This immature, proliferative state creates a vulnerable cell population prone to oncogenic transformation, especially in a microenvironment altered by MEN1 loss. Targeting INSM1 and its associated pathways could help prevent the transition from molecular dysregulation to overt gastrinoma, thereby reducing the burden of complications like Zollinger-Ellison syndrome.

INSM1 is not only a marker of neuroendocrine identity but a mechanistic mediator of tumor predisposition in MEN1-deficient patients. Its early detection through molecular diagnostics, coupled with its potential for targeted modulation, positions INSM1 as a critical node in precision surveillance, early diagnosis, and future therapeutic design for gastrinomas. Incorporating INSM1 into MEN1 surveillance protocols could shift the clinical paradigm from reactive intervention to preemptive, personalized management, substantially improving outcomes in this high-risk patient population.

## 8. ARX

### Mutated MEN1 in Dysregulating Enteroendocrine Cells

ARX (Aristaless-related homeobox) is a key transcription factor involved in the differentiation and function of neuroendocrine cells, including those in the duodenum. In the context of enteroendocrine cells, ARX plays a crucial role in controlling the fate of progenitor cells and their differentiation into specific hormone-producing cell types. In the normal developmental process, ARX is required for the establishment of a functional enteroendocrine cell population, including cells that secrete important hormones such as somatostatin and glucagon-like peptide 1 (GLP-1). This transcription factor is tightly regulated by a variety of signaling pathways, including those mediated by MEN1, which encodes menin, a tumor suppressor protein that regulates multiple transcription factors involved in cell fate decisions [65]. Menin is known to form complexes with various transcription factors, acting as a cofactor to regulate gene expression, including that of ARX. The loss of MEN1 disrupts this regulatory process, leading to potential dysregulation of ARX expression, which could impair the differentiation and function of neuroendocrine cells in the duodenum. When MEN1 is lost, the loss of menin’s regulatory role disrupts ARX gene expression in multiple ways. In particular, without proper menin-mediated regulation, ARX expression may become either insufficient or excessively expressed in progenitor cells. A reduction in ARX levels may lead to incomplete differentiation of enteroendocrine cells, resulting in a failure to produce fully functional hormone-secreting cells. Conversely, overexpression of ARX may lead to the proliferation of undifferentiated cells or the formation of a disorganized population of hormone-secreting cells. This disruption in ARX expression and function can hinder the delicate balance between progenitor cell proliferation and differentiation, a balance essential for maintaining homeostasis within the enteroendocrine compartment of the duodenum [66]. The dysregulation of ARX expression due to MEN1 loss can have profound consequences on enteroendocrine cell lineage specification. ARX is known to interact with other transcription factors such as ISL1 and PAX4, which are also essential for the proper differentiation of enteroendocrine cells. The loss of MEN1 may disrupt the coordinated expression of these transcription factors, further impairing cell fate decisions in the duodenum. This could result in an expansion of less differentiated progenitor cells or a failure to produce specific neuroendocrine cell types, ultimately leading to an altered hormone production profile within the duodenum. In particular, the lack of fully differentiated enteroendocrine cells could contribute to a dysfunctional regulatory environment, which may predispose to tumorigenesis, including neuroendocrine tumors like gastrinomas.

Additionally, the impact of MEN1 loss on ARX may disrupt the normal feedback mechanisms that regulate the growth and function of enteroendocrine cells. In the absence of menin, ARX may lose its ability to appropriately respond to external signals such as Notch or Wnt signaling, which are crucial for the maintenance of proper progenitor cell pools and their differentiation into mature cell types [67]. This disruption of ARX regulation could lead to an imbalance between self-renewing progenitors and differentiated cells, resulting in a proliferative state that favors tumorigenesis. Over time, such dysregulation could promote the formation of gastrinomas or other types of neuroendocrine tumors in the duodenum, as the loss of control over cellular differentiation and proliferation creates a favorable environment for malignant transformation. This loss of MEN1 can lead to dysregulation of ARX, a critical transcription factor in the development and differentiation of neuroendocrine cells in the duodenum. Without proper MEN1-mediated regulation, ARX expression can become aberrant, leading to incomplete or excessive differentiation of enteroendocrine cells. This disruption impairs the balance between cell proliferation and differentiation, which is essential for maintaining normal hormone secretion and intestinal function. The resulting abnormal population of neuroendocrine cells, coupled with potential disruptions in other signaling pathways, increases the risk of tumorigenesis, particularly the development of gastrinomas, which are associated with overproduction of gastric acid. The loss of MEN1 thus plays a central role in the dysregulation of ARX and the cellular processes that drive neuroendocrine tumor formation in the duodenum [68].

### Consequences for Enteroendocrine Cells and Associated Gastrionoma Risk

The loss of MEN1 has significant implications for the development and differentiation of neuroendocrine cells in the duodenum, particularly through its effect on the transcription factor ARX. ARX plays an essential role in the proper differentiation and function of enteroendocrine cells, which produce key hormones involved in the regulation of gastrointestinal physiology. Under normal conditions, ARX is tightly regulated, helping maintain the delicate balance between progenitor cell proliferation and differentiation into hormone-secreting cell types. Menin, the protein encoded by the MEN1 gene, serves as a critical regulator of transcriptional networks, including those that control ARX expression [69]. When MEN1 is lost, the regulatory control over ARX is disrupted, which can lead to abnormal expression patterns and function of ARX in the duodenal neuroendocrine cells. In the absence of MEN1, ARX expression may become either insufficient or excessively activated. If ARX is underexpressed due to the loss of MEN1, enteroendocrine progenitor cells may fail to fully differentiate into the appropriate mature hormone-producing cells, leading to a deficiency in necessary hormones such as somatostatin and glucagon-like peptide-1 (GLP-1). This incomplete differentiation can result in a disruption of normal gastrointestinal function and cellular homeostasis. On the other hand, if ARX is overexpressed, progenitor cells may fail to undergo proper differentiation and may instead proliferate in an uncontrolled manner, expanding the pool of undifferentiated cells or producing abnormal cell types. This unregulated proliferation could increase the risk of tumorigenesis, including the development of gastrinomas, a type of neuroendocrine tumor. Furthermore, ARX is known to interact with other transcription factors such as ISL1, PAX4, and PAX6, which coordinate the differentiation of specific enteroendocrine cell types. The loss of MEN1 may disrupt the interplay between these transcription factors, further exacerbating the dysregulation of ARX expression. This disruption can alter the fate of enteroendocrine cells, pushing them toward aberrant cell fates that promote tumorigenic growth [70]. Additionally, without the regulatory function of menin, ARX may fail to respond appropriately to critical signaling pathways such as Notch and Wnt, which are important for maintaining the balance between progenitor cell proliferation and differentiation. The dysregulated ARX signaling in this context can lead to an unchecked expansion of progenitor cells or a failure to generate differentiated neuroendocrine cells, creating a microenvironment conducive to the development of gastrinomas. The loss of MEN1 and its effect on ARX expression ultimately disrupts the normal function and differentiation of enteroendocrine cells, increasing the likelihood of tumor formation. The failure to maintain appropriate regulation of progenitor cell proliferation and differentiation contributes to a pathophysiological state where the risk of gastrinomas and other neuroendocrine tumors in the duodenum is significantly heightened. The combination of abnormal ARX expression and impaired signaling pathways that control cell fate decisions plays a central role in the tumorigenic process, highlighting the critical role of MEN1 in maintaining the integrity of neuroendocrine differentiation and preventing the onset of malignancies like gastrinomas [71, 72].

### Possible Clinical Implications of ARX Dysregulation in MEN1 Patients: Enhancing Diagnostic Precision, Risk Stratification, and Therapeutic Targeting in Gastrinoma Prevention

ARX is a central transcription factor involved in specifying the fate of neuroendocrine progenitor cells in the gastrointestinal tract, particularly influencing the differentiation of hormone-producing enteroendocrine cells. In the duodenum, ARX cooperates with other key regulators such as PAX4, ISL1, and PAX6 to direct the balance between progenitor maintenance and differentiation into functional hormone-secreting cells. The loss of MEN1, which encodes the tumor suppressor menin, impairs the regulation of ARX at the epigenetic and transcriptional levels, leading to its aberrant expression and increasing susceptibility to neuroendocrine tumor formation, including gastrinomas. From a clinical standpoint, the dysregulation of ARX represents a critical biomarker of disrupted endocrine lineage specification, which can be detected early in MEN1 patients through biopsy-based molecular profiling. ARX expression levels either insufficient or excessively high may signal a deviation from normal differentiation programs.

Incorporating ARX immunohistochemistry or transcript analysis into duodenal biopsy surveillance during routine endoscopy could improve early risk identification, even in the absence of visible lesions, enabling a shift toward molecularly informed screening and surveillance protocols. Moreover, aberrant ARX activity contributes to the expansion of undifferentiated or mis-specified enteroendocrine cells, particularly those biased toward gastrin production. This imbalance can increase the likelihood of gastrin-producing progenitor cell proliferation, creating a substrate for gastrinoma formation. As such, ARX profiling especially when combined with expression data for NEUROG3, PAX4, and INSM may help stratify MEN1 patients into molecular risk tiers, guiding decisions around surveillance intensity and timing of intervention.

Therapeutically, ARX dysregulation suggests a potential role for differentiation-restoring strategies. In cases of overexpression, targeting upstream pathways such as Notch, Wnt, or chromatin remodeling machinery could help re-establish the regulatory feedback needed to enforce proper progenitor maturation. Conversely, in cases of ARX underexpression, epigenetic therapies that reactivate silenced differentiation programs may reduce the persistence of immature cell pools. Either approach could contribute to chemoprevention strategies in high-risk MEN1 carriers. Importantly, ARX’s function in maintaining neuroendocrine identity and regulating hormonal output highlights its relevance not only for tumor initiation but also for the functional outcomes of tumor biology. Aberrant ARX activity may lead to dysregulated gastrin production and associated complications such as Zollinger-Ellison syndrome.

Therefore, ARX serves as a dual-purpose target: both for early tumor risk detection and for understanding and mitigating functional consequences of hormone overproduction in MEN1-associated gastrinomas. The dysregulation of ARX following MEN1 loss plays a mechanistic and clinically actionable role in the initiation of duodenal neuroendocrine tumors. Monitoring ARX expression in biopsy specimens, combined with other neuroendocrine transcriptional markers, offers an opportunity to personalize surveillance, enable earlier diagnosis, and inform future therapies that aim to correct the underlying differentiation defects driving tumorigenesis. ARX thus stands as a vital node in the precision management framework for MEN1-related neuroendocrine neoplasia.

## 9. Notch Pathway

### Mutated MEN1 in Dysregulating Enteroendocrine Cells

The Notch signaling pathway plays a crucial role in the development and differentiation of neuroendocrine (enteroendocrine) cells in the duodenum by regulating the balance between progenitor cell proliferation and differentiation. Notch signaling is known to maintain the undifferentiated state of progenitor cells, and its activation promotes the commitment of these cells to specific fates [73]. It works through a series of interactions involving Notch receptors and ligands, which ultimately control the expression of downstream target genes that influence cell fate decisions. In the context of MEN1, the loss of meni a critical tumor suppressor encoded by the MEN1 gene disrupts the proper regulation of this pathway, potentially leading to abnormal cellular outcomes that can predispose to tumorigenesis. Menin is known to interact with various transcription factors and chromatin regulators to maintain proper signaling pathways, including Notch. The loss of MEN1 leads to a lack of control over this interaction, which could result in either hyperactivation or insufficient activation of the Notch pathway in enteroendocrine progenitor cells. In particular, loss of menin can lead to the accumulation of Notch ligands or an increased sensitivity of the Notch receptors, resulting in excessive activation of the pathway [74]. This overactivation may push progenitor cells into an undifferentiated state, preventing them from properly differentiating into mature enteroendocrine cells. Alternatively, insufficient activation of Notch signaling, which could occur through the failure of menin to appropriately regulate this pathway, may lead to premature differentiation of progenitor cells. This imbalance in the Notch signaling cascade would disrupt normal lineage commitment, leading to aberrant differentiation patterns in the duodenum. The disruption of Notch signaling due to the loss of MEN1 could have serious implications for the development and function of enteroendocrine cells, which rely on finely tuned signaling for proper differentiation into hormone-producing cells such as somatostatin-and glucagon-like peptide-1 (GLP-1)-secreting cells. If Notch signaling is dysregulated, this can impair the formation of these mature cell types, resulting in a deficit of critical hormones required for gastrointestinal function and homeostasis [75]. Furthermore, the failure of progenitor cells to undergo proper differentiation could contribute to the unchecked proliferation of undifferentiated or partially differentiated cells, increasing the risk of tumor formation, such as gastrinomas, a common neuroendocrine tumor associated with MEN1 loss. Additionally, the loss of MEN1 may affect the interaction between Notch signaling and other key regulatory pathways involved in neuroendocrine differentiation, such as Wnt and Notch-related transcription factors like Hes1, which are also regulated by menin. These alterations could lead to an expansion of progenitor cell populations, a failure of normal differentiation, or a shift toward a more proliferative, tumor-prone phenotype. The lack of proper differentiation due to Notch pathway dysregulation could ultimately set the stage for the development of gastrinomas in the duodenum. As progenitor cells accumulate and fail to differentiate into specialized neuroendocrine cells, the risk of tumorigenesis escalates, driven by the disruption in the delicate balance of cellular differentiation and proliferation that Notch signaling normally helps to maintain. The loss of MEN1 has profound consequences for Notch signaling in enteroendocrine cells in the duodenum. By disrupting the regulation of this pathway, the loss of MEN1 can either cause the overproliferation of progenitor cells or promote premature differentiation, both of which can contribute to the pathological development of neuroendocrine tumors like gastrinomas. The inability to properly control the balance between differentiation and proliferation underlines the critical role of MEN1 in maintaining normal gastrointestinal homeostasis and preventing tumorigenesis in the duodenum [76].

### Consequences for Enteroendocrine Cells and Associated Gastrionoma Risk

The loss of MEN1 (Multiple Endocrine Neoplasia type 1) can significantly disrupt the Notch signaling pathway in enteroendocrine cells of the duodenum, which plays a critical role in regulating cell fate determination and differentiation. Notch signaling in these cells is essential for maintaining a balance between progenitor cell proliferation and differentiation into mature, functional neuroendocrine cells. Notch activation promotes the undifferentiated state of progenitor cells and prevents them from prematurely differentiating. Under normal circumstances, MEN1 interacts with various transcription factors and chromatin regulators, including those controlling the Notch pathway, to ensure appropriate signaling. When MEN1 is lost, this regulatory function is compromised, leading to dysregulated Notch signaling that can disrupt the normal differentiation of neuroendocrine cells and increase the risk of tumorigenesis, particularly gastrinomas [77]. In the absence of MEN1, Notch signaling may become either hyperactive or insufficiently activated, leading to an imbalance in the differentiation process. Hyperactivation of Notch signaling in progenitor cells can prevent them from differentiating into their intended mature cell types, such as the hormone-secreting enteroendocrine cells. This results in an accumulation of undifferentiated or partially differentiated cells that continue to proliferate rather than specialize. In this scenario, the progenitor cells are kept in an immature state, which is a hallmark of tumorigenesis. On the other hand, insufficient Notch activation may lead to premature differentiation of progenitor cells, preventing the normal pool of progenitor cells from expanding. This premature differentiation can limit the number of functional neuroendocrine cells and also lead to a defective hormone-producing capacity, further contributing to the tumorigenic environment. This disruption of Notch signaling under MEN1 loss could lead to an increased risk of gastrinoma formation in the duodenum.

Gastrinomas are a type of neuroendocrine tumor that arises from enteroendocrine cells, particularly those that secrete gastrin. The failure to properly differentiate into mature enteroendocrine cells due to dysregulated Notch signaling can result in a tumorigenic microenvironment where proliferating cells are more likely to acquire mutations that drive cancerous growth [78]. These undifferentiated or partially differentiated cells, which persist due to the improper regulation of Notch, could acquire additional genetic mutations that enable them to form gastrinomas. Moreover, the failure of appropriate differentiation may lead to an overproduction of certain hormones, such as gastrin, which is a defining feature of gastrinomas and contributes to the tumor’s ability to grow and metastasize. The dysregulation of Notch signaling in the context of MEN1 loss not only compromises the differentiation of enteroendocrine cells but also disrupts the fine-tuned regulation required for normal gastrointestinal function. This imbalance creates an environment where the progenitor cells are more likely to undergo abnormal expansion, fail to differentiate, and eventually progress toward a cancerous phenotype. As a result, the loss of MEN1, through its effect on Notch signaling, plays a key role in increasing the risk of gastrinomas by promoting the proliferation of undifferentiated neuroendocrine cells that are predisposed to malignant transformation [79, 80].

### Possible Clinical Implications of Notch Pathway Dysregulation in MEN1 Patients: Enhancing Early Diagnosis, Risk Stratification, and Therapeutic Intervention in Gastrinoma

The Notch signaling pathway plays a central role in the fate determination of duodenal enteroendocrine progenitors, regulating the delicate balance between proliferation and differentiation. In healthy tissue, Notch maintains a progenitor state by preventing premature differentiation; its modulation is essential for ensuring appropriate endocrine lineage maturation. However, in the context of MEN1 loss, the regulatory scaffold protein menin is absent, leading to either aberrant activation or suppression of Notch signaling. This disruption skews the differentiation landscape and creates a pro-tumorigenic environment, particularly predisposing to gastrin-secreting neuroendocrine tumors (gastrinomas). Clinically, Notch dysregulation is an early, actionable event in the trajectory of neuroendocrine tumorigenesis. MEN1 patients undergoing routine duodenal biopsies as part of surveillance protocols may benefit from Notch pathway profiling, including assessment of Notch receptor expression, ligand levels (e.g., DLL1, Jagged), and downstream targets such as Hes1. Aberrant pattern such as overexpression of Hes1 or suppressed differentiation markers could serve as early molecular indicators of progenitor cell arrest, preceding visible tumor formation. These insights support biopsy-based diagnostics aimed at identifying patients at high molecular risk of gastrinoma.

From a surveillance perspective, the presence of Notch pathway dysregulation especially when correlated with reduced expression of downstream differentiation factors (e.g., NEUROG3, PAX6, or ISL1) can stratify MEN1 patients into risk tiers. This enables precision surveillance, in which patients with sustained Notch activation and proliferative markers undergo more frequent endoscopic evaluations and molecular re-biopsies, while those with balanced differentiation signals may follow standard protocols. Therapeutically, the Notch pathway is a viable target for intervention. Preclinical studies and early-phase clinical trials have explored Notch inhibitors and modulators (e.g., γ-secretase inhibitors) for controlling tumor growth in various neuroendocrine and solid tumors. In the context of MEN1-related gastrinoma, therapeutic Notch modulation could help redirect progenitor cells toward terminal differentiation, reducing the reservoir of undifferentiated, mutation-prone cells. This approach could form the basis of chemopreventive or differentiation-restoring therapy in MEN1 patients identified as high-risk by molecular screening.

Furthermore, since Notch signaling intersects with other critical pathways disrupted by MEN1 loss such as Wnt, MAPK, and epigenetic regulators, the integration of Notch modulation into multi-targeted therapy regimens may yield more effective results. This is particularly relevant for gastrinoma prevention, where the pathogenesis involves both aberrant cell proliferation and unregulated gastrin secretion.

## 10. WNT Pathway

### Mutated MEN1 in Dysregulating Enteroendocrine Cells

The Wnt signaling pathway plays an important role in regulating the development and differentiation of neuroendocrine cells, including those in the duodenum. In enteroendocrine cells, Wnt signaling is involved in controlling cell fate decisions, maintaining stem cell populations, and influencing the differentiation of progenitor cells into specialized hormone-secreting cells. When functioning properly, the Wnt pathway helps maintain a balance between cell proliferation and differentiation, ensuring proper development of the enteroendocrine cell lineage.

MEN1, a tumor suppressor gene, is known to be involved in regulating various cellular processes, including those governing transcriptional networks that control Wnt signaling [81]. The loss of MEN1 can disrupt the delicate regulation of Wnt signaling, leading to aberrant gene expression that may have serious consequences for the development and function of enteroendocrine cells in the duodenum. Under normal conditions, MEN1 is involved in chromatin remodeling and transcriptional regulation, which influences the expression of key components within the Wnt signaling pathway. MEN1 may help modulate the activity of several transcription factors that regulate the Wnt pathway, ensuring that Wnt signals are activated or repressed appropriately during the development of neuroendocrine cells. When MEN1 is lost, this regulatory function is compromised, which can lead to the dysregulation of Wnt signaling. This dysregulation can manifest in two ways. First, the pathway could become constitutively activated, leading to excessive Wnt signaling. Overactive Wnt signaling in progenitor cells and early-stage enteroendocrine cells may result in abnormal cell proliferation, preventing proper differentiation and causing the accumulation of undifferentiated cells. These cells may be more prone to tumorigenic transformation due to their uncontrolled proliferation. Second, the loss of MEN1 could lead to insufficient Wnt signaling, which could hinder the expansion of stem and progenitor cells . This would disrupt the normal differentiation process, leading to a deficiency of mature enteroendocrine cells, potentially impairing the normal function of the duodenum and contributing to the development of tumors such as gastrinomas. The loss of MEN1-induced dysregulation of Wnt signaling in enteroendocrine cells of the duodenum can also affect key pathways that control the balance between stem cell renewal and differentiation [82]. Wnt signaling is known to regulate the expression of several critical genes that control progenitor cell maintenance and differentiation into hormone-producing cells. When MEN1 is lost, the resulting alterations in Wnt signaling could lead to the inappropriate activation of genes that promote cell proliferation, such as c-myc or cyclin D1. Alternatively, insufficient Wnt signaling could prevent the activation of genes that promote differentiation, such as neurogenin and other transcription factors involved in neuroendocrine differentiation. These disruptions can impair the ability of progenitor cells to properly differentiate into specialized enteroendocrine cells, leading to an accumulation of undifferentiated cells that can undergo malignant transformation. Furthermore, the dysregulation of Wnt signaling due to MEN1 loss may alter the molecular interactions between Wnt and other signaling pathways, such as Notch and BMP, that are also crucial for neuroendocrine cell differentiation [83]. The interplay between these pathways is critical for maintaining the proper differentiation trajectory of progenitor cells, and disruptions in this balance can further promote abnormal growth and differentiation patterns, leading to the formation of neuroendocrine tumors, including gastrinomas. As a result, the loss of MEN1 disrupts the finely tuned regulation of Wnt signaling, which can lead to an imbalance between cell proliferation and differentiation, increasing the risk of tumorigenesis and the development of gastrinomas in the duodenum [84].

### Consequences for Enteroendocrine Cells and Associated Gastrionoma Risk

The loss of MEN1 can significantly impact the Wnt signaling pathway, leading to disruptions in the development and differentiation of neuroendocrine (enteroendocrine) cells in the duodenum. Wnt signaling is a key regulator of cell fate decisions, proliferation, and differentiation in a variety of tissues, including the gut. In enteroendocrine cells, Wnt signaling helps control the balance between stem cell maintenance and differentiation into specialized hormone-secreting cells. The proper regulation of Wnt signaling is crucial for ensuring that progenitor cells differentiate into the correct cell types and maintain normal function. MEN1, a tumor suppressor gene, plays a pivotal role in regulating several cellular processes, including chromatin remodeling and transcriptional control. It affects the expression of key components in signaling pathways, such as Wnt, by directly influencing transcription factors that control these pathways [85]. When MEN1 is lost, its regulatory functions are compromised, leading to the dysregulation of Wnt signaling. One potential consequence of MEN1 loss is the constitutive activation of the Wnt pathway. In this scenario, the increased activation of Wnt signaling may result in excessive proliferation of progenitor cells, including those within the duodenal neuroendocrine lineage. This uncontrolled cell division prevents proper differentiation into mature enteroendocrine cells, causing an accumulation of undifferentiated cells that are more likely to undergo malignant transformation. These proliferating progenitor cells may acquire additional mutations or alterations in gene expression, further promoting tumorigenesis. Overactive Wnt signaling can also prevent the differentiation of progenitor cells into the appropriate neuroendocrine cell types, which impairs the function of the duodenum and may contribute to the development of gastrinomas, a type of neuroendocrine tumor that arises from cells producing gastrin [86]. On the other hand, loss of MEN1 could also result in insufficient activation of the Wnt pathway, which would impair the maintenance of stem cells and progenitor cells in the duodenum. Inadequate Wnt signaling can hinder the differentiation of these progenitor cells into mature enteroendocrine cells, leading to a shortage of functional hormone-secreting cells. This imbalance in cell fate decisions may not only disrupt normal gut physiology but also increase the likelihood of tumorigenesis. Insufficient Wnt signaling could lead to a dysregulated differentiation process in which cells fail to acquire the appropriate functional identity, increasing the risk of abnormal cell growth and the development of neuroendocrine tumors such as gastrinomas. Additionally, MEN1’s loss may affect the interactions between the Wnt pathway and other signaling pathways, such as Notch and BMP, which are also involved in controlling the fate of enteroendocrine cells. These pathways often work together to maintain proper cellular balance between self-renewal, proliferation, and differentiation [87]. When MEN1 is lost and Wnt signaling becomes disrupted, the cross-talk between these pathways may also be altered, further exacerbating the effects on cell fate determination and contributing to the development of gastrinomas. The resultant disruption in both Wnt signaling and its downstream effects on enteroendocrine cell differentiation ultimately increases the risk of tumor formation in the duodenum, especially in the form of gastrinomas, which are characterized by abnormal gastrin-producing cells. Therefore, the loss of MEN1 has the potential to disrupt Wnt-mediated regulation of neuroendocrine cell fate, leading to an increased likelihood of tumorigenesis in the duodenum [88].

### Possible Clinical Implications of Wnt Pathway Dysregulation in MEN1 Patients: Advancing Early Detection, Risk Stratification, and Therapeutic Precision in Gastrinoma

The Wnt signaling pathway plays a fundamental role in orchestrating the balance between stem cell maintenance, progenitor expansion, and differentiation of enteroendocrine cells in the gastrointestinal tract. In normal duodenal development, Wnt tightly regulates the fate of neuroendocrine precursors to ensure appropriate maturation into specialized hormone-producing cells, including gastrin-secreting G cells. In MEN1-deficient states, the loss of menin impairs chromatin regulation and transcriptional control, leading to Wnt pathway dysregulation, an important event in the pathogenesis of gastrinomas.

From a clinical standpoint, Wnt dysregulation represents a valuable molecular marker of early endocrine cell misprogramming. Assessing Wnt pathway activation status via markers such as β-catenin nuclear localization, c-Myc, or Cyclin D1 expression in duodenal biopsy specimens can offer early insight into neoplastic risk in MEN1 patients.

This approach enables clinicians to move beyond conventional imaging and gastrin-level monitoring toward molecularly guided surveillance. The presence of Wnt activation or suppression signatures, even in morphologically normal mucosa, could serve as a red flag for disrupted progenitor differentiation, prompting intensified follow-up and preventative planning. Furthermore, Wnt pathway status can serve as a stratification tool in MEN1 patients.

Overactivation of Wnt signaling suggests excessive progenitor proliferation and impaired differentiation, raising the likelihood of tumor-initiating cell pools. Underactivation of Wnt may indicate inadequate progenitor maintenance, leading to dysfunctional or misdifferentiated cells that can accumulate genetic instability over time. These molecular subtypes can guide personalized surveillance frequency, particularly when integrated with expression data from other MEN1-affected regulators (e.g., NEUROG3, INSM1, PAX4/6).

Therapeutically, the Wnt pathway presents a tractable target. In patients showing Wnt hyperactivation, Wnt inhibitors (e.g., PORCN inhibitors, β-catenin antagonists) may curb progenitor overexpansion and reestablish proper lineage restriction. Alternatively, in cases of Wnt underactivity, agents that restore Wnt signaling balance could support normal differentiation and reduce the persistence of undifferentiated, transformation-prone cells. These differentiation-based strategies have growing support in the management of early neuroendocrine neoplasia and may be particularly beneficial in MEN1 mutation carriers with molecular evidence of early dysregulation. Importantly, Wnt signaling intersects with other dysregulated pathways in MEN1-deficient tissues especially Notch and BMP, both of which also govern neuroendocrine differentiation. Cross-talk between these pathways is essential for maintaining cellular balance, and MEN1 loss disrupts this interaction. Therefore, multi-pathway modulation strategies may offer synergistic benefits in restoring proper cell fate dynamics and reducing the risk of tumorigenesis.

Monitoring Wnt activity in biopsy-based diagnostics, using it as a molecular risk stratifier, and targeting it pharmacologically can significantly improve early detection, surveillance precision, and therapeutic intervention in MEN1 patients at risk for gastrinoma.

## 11. BMP Pathway

### Mutated MEN1 in Dysregulating Enteroendocrine Cells

The Bone Morphogenetic Protein (BMP) signaling pathway plays a critical role in regulating the differentiation and development of various cell types, including neuroendocrine (enteroendocrine) cells in the duodenum. BMPs are involved in controlling cellular processes such as proliferation, apoptosis, and lineage specification, and they help guide the proper differentiation of progenitor cells into specialized cell types. In the context of neuroendocrine cells, BMP signaling is essential for maintaining a balance between stem cell proliferation and differentiation [89]. The transcription factors regulated by BMP signaling contribute to determining cell fate, ensuring that progenitor cells differentiate into functional enteroendocrine cells capable of producing hormones like gastrin, secretin, and somatostatin. The Menin protein, encoded by the MEN1 gene, plays a vital role in regulating transcriptional processes and in the proper function of various signaling pathways, including BMP signaling. Loss of MEN1 disrupts the regulatory network that governs BMP signaling, leading to the potential dysregulation of this pathway in neuroendocrine cells of the duodenum. When MEN1 is lost, the normal control over BMP signaling is compromised, which can lead to either excessive or insufficient activation of the BMP pathway in duodenal neuroendocrine cells. A dysregulated BMP pathway can result in abnormal cell fate decisions, which in turn can disturb the proper differentiation of progenitor cells into mature enteroendocrine cells. Overactivation of BMP signaling due to MEN1 loss may lead to an increase in apoptosis or a failure to properly differentiate progenitor cells into functional hormone-secreting cells [90]. This can cause an imbalance in the duodenal cell population, with a disproportionate number of undifferentiated progenitor cells, or the development of cells that are functionally abnormal. Furthermore, dysregulated BMP signaling can alter the cross-talk between BMP and other developmental pathways, such as Notch and Wnt, which further exacerbates the disruption in differentiation. These molecular disruptions in cell fate determination can increase the risk of aberrant growth and the potential for neoplastic transformation, ultimately contributing to the formation of tumors like gastrinomas in the duodenum. On the other hand, insufficient BMP signaling caused by MEN1 loss may prevent the proper differentiation of progenitor cells into enteroendocrine cells, resulting in a shortage of hormone-producing cells in the duodenum. This insufficient differentiation could lead to defective intestinal functions, as the loss of these important hormone-secreting cells would disrupt the regulation of digestive processes. In the context of neuroendocrine cell development, a reduction in BMP signaling can also impair the signaling network necessary for maintaining stem cell populations, leading to an inability to replenish the enteroendocrine cell lineage. This imbalance between stem cell renewal and differentiation may further promote tumorigenesis, particularly in the form of gastrinomas, as the dysregulated differentiation processes in progenitor cells could result in the accumulation of cells prone to malignant transformation. Moreover, the loss of MEN1 could alter the delicate temporal and spatial regulation of BMP signaling in the duodenum. BMPs need to be precisely regulated to ensure the correct timing of differentiation [91]. MEN1, through its regulation of transcription and chromatin remodeling, is integral to this precise control. Without MEN1, the BMP pathway may become chronically active or insufficiently activated, disrupting the normal developmental progression of enteroendocrine cells. Such dysregulation of BMP signaling in the absence of MEN1 could therefore lead to an increased susceptibility to the formation of abnormal neuroendocrine tumors, such as gastrinomas, driven by improperly differentiated or unregulated enteroendocrine cells. Ultimately, the loss of MEN1 interferes with BMP signaling, leading to a disruption in the developmental biology of neuroendocrine cells in the duodenum and increasing the likelihood of neoplastic growth [92].

### Consequences for Enteroendocrine Cells and Associated Gastrionoma Risk

The loss of MEN1, a gene that encodes the tumor suppressor protein Menin, has a significant impact on various signaling pathways, including BMP (Bone Morphogenetic Protein) signaling, which plays a crucial role in regulating the differentiation of neuroendocrine cells in the duodenum. BMPs are essential for a variety of cellular processes, including cell proliferation, differentiation, and apoptosis, and they are involved in maintaining the balance between stem cell renewal and differentiation. In neuroendocrine cells, BMP signaling helps regulate the differentiation of progenitor cells into functional hormone-producing enteroendocrine cells, which are responsible for secreting important regulatory hormones such as gastrin. The loss of MEN1 disrupts this regulatory process by altering BMP signaling, which can lead to either excessive or insufficient activation of the pathway, both of which have implications for cell fate decisions in the duodenum. When MEN1 is lost, the normal control over BMP signaling is compromised, which can lead to dysregulation of the differentiation of neuroendocrine progenitor cells. One consequence of this is the potential failure to properly differentiate into mature enteroendocrine cells. Dysregulated BMP signaling can result in progenitor cells failing to commit to a neuroendocrine fate, or conversely, it could drive progenitors to differentiate into cells with an altered function [93]. This improper differentiation can lead to an overabundance of cells that are either undifferentiated or abnormally differentiated, which could disrupt the balance of hormone production in the duodenum. Additionally, the loss of MEN1 may prevent the timely differentiation of these cells, leading to the accumulation of progenitor cells that are more likely to undergo malignant transformation. Such transformation could result in tumorigenesis, particularly in the form of gastrinomas, as the disordered differentiation of cells creates a favorable environment for oncogenic alterations. Furthermore, when BMP signaling is dysregulated due to the loss of MEN1, it can interfere with the interaction between BMP and other signaling pathways, such as Notch and Wnt, which also contribute to neuroendocrine cell fate decisions. This cross-talk between signaling pathways is vital for ensuring the proper timing and spatial arrangement of cell differentiation. Disruption of BMP signaling due to MEN1 loss could lead to aberrant cross-talk between these pathways, which would further exacerbate the imbalance in the differentiation of neuroendocrine cells [94]. This dysregulation could result in abnormal growth patterns of neuroendocrine cells, potentially fostering an environment in which cells are more prone to malignant transformation and neoplastic growth. In addition to its role in differentiation, BMP signaling also influences stem cell behavior, and the loss of MEN1 could result in an inability to maintain the normal population of enteroendocrine progenitor cells. This failure to regulate the balance between stem cell renewal and differentiation could lead to the depletion of functional enteroendocrine cells, further exacerbating the risk of tumorigenesis. The altered behavior of progenitor cells in the absence of MEN1 could promote the expansion of undifferentiated or abnormally differentiated cells, which in turn increases the likelihood of these cells undergoing malignant transformation. This malignant transformation, driven by the loss of MEN1 and dysregulated BMP signaling, could result in the development of tumors such as gastrinomas, which are characterized by excessive production of gastrin and other hormones by neuroendocrine cells. Overall, the loss of MEN1 disrupts the normal regulation of BMP signaling in neuroendocrine cells in the duodenum, which has profound consequences for cell fate determination. The altered BMP pathway can lead to improper differentiation of progenitor cells, an imbalance in the number and function of enteroendocrine cells, and an increased risk of neoplastic transformation. The dysregulated differentiation of cells in the absence of MEN1, particularly in the context of abnormal BMP signaling, contributes to the formation of tumors such as gastrinomas, highlighting the critical role that MEN1 plays in maintaining the proper function of neuroendocrine cells in the duodenum and preventing tumorigenesis [95, 96].

### Possible Clinical Implications of BMP Pathway Dysregulation in MEN1 Patients: Advancing Early Detection, Risk Stratification, and Therapeutic Targeting of Gastrinoma

The BMP signaling pathway plays a critical role in directing the differentiation, apoptosis, and lineage specification of duodenal neuroendocrine progenitors into mature hormone-producing enteroendocrine cells. The loss of MEN1, a tumor suppressor gene encoding menin, disrupts transcriptional and epigenetic regulation of BMP signaling, leading to impaired differentiation dynamics and increased tumor susceptibility, particularly in the form of gastrinomas.

Clinically, this insight positions BMP dysregulation as a valuable molecular biomarker in early-stage disease surveillance for MEN1 mutation carriers. Duodenal biopsy specimens collected during routine endoscopic monitoring can be analyzed for BMP pathway activity, such as levels of phospho-SMAD1/5/8, BMP ligands (e.g., BMP4, BMP7), or downstream differentiation targets. A reduction or aberrant elevation of BMP signaling may indicate an imbalance between progenitor maintenance and differentiation, a hallmark of pre-neoplastic transformation. This makes BMP signaling assessment a promising tool for early detection of molecular dysregulation prior to histologically visible tumor formation. In terms of risk stratification, BMP signaling status may help define which MEN1 patients are most likely to progress from molecular dysregulation to overt neuroendocrine tumorigenesis.

Overactivation of BMP signaling could suppress neuroendocrine differentiation, increase apoptosis, or prevent proper maturation leading to accumulation of dysfunctional or stressed progenitor cells prone to transformation. Insufficient BMP activity may result in failure of proper lineage commitment, permitting a buildup of undifferentiated, proliferative precursors that fuel gastrinoma initiation. Surveillance protocols could be customized based on BMP activity, allowing for closer monitoring of high-risk patients and potentially earlier therapeutic intervention in those showing early dysregulation.

From a therapeutic perspective, BMP dysregulation opens opportunities for differentiation therapy or signaling pathway modulation. Agents that restore BMP balance whether through recombinant BMPs, BMP receptor agonists/antagonists, or SMAD-targeted modulation could support proper endocrine cell maturation and reduce the persistence of transformation-prone progenitors. Moreover, epigenetic therapies that restore menin-mediated chromatin control may re-establish BMP pathway integrity. Additionally, BMP signaling does not operate in isolation. Its interaction with Notch, Wnt, and MAPK pathways, all of which are also disrupted by MEN1 loss, means that BMP-targeted approaches could be enhanced by multi-pathway therapy. Understanding this cross-talk could help clinicians design combinatorial treatments to reinforce differentiation signals and suppress tumor-initiating cell populations in MEN1 patients. Importantly, BMP signaling influences not only tumor risk but also duodenal hormonal function. BMP dysregulation may skew the endocrine cell pool, leading to abnormal gastrin production and contributing to clinical syndromes such as Zollinger-Ellison syndrome even in the absence of overt tumor burden. Thus, early detection and correction of BMP imbalances could also alleviate functional disease manifestations.

## 12. Shh Pathway

### Mutated MEN1 in Dysregulating Enteroendocrine Cells

The Sonic Hedgehog (Shh) signaling pathway plays a critical role in the development of various tissues, including the differentiation of neuroendocrine (enteroendocrine) cells in the duodenum. Shh is a key regulator of progenitor cell fate, influencing cellular processes such as proliferation, differentiation, and patterning during early development. In the context of neuroendocrine cells, Shh is involved in maintaining the balance between stem cell renewal and differentiation, ensuring the proper development of specialized hormone-secreting cells in the gut. The loss of MEN1, a gene encoding the Menin protein, can disrupt this delicate balance by altering the regulation of Shh signaling, potentially leading to dysregulated cell differentiation and an increased risk of pathological conditions such as gastrinomas [97]. MEN1 plays an essential role in regulating various signaling pathways, including Shh, by modulating the expression of target genes and interacting with transcription factors that are involved in cell differentiation and proliferation. The loss of MEN1 results in the loss of its regulatory control over these pathways, which may lead to aberrant activation or inhibition of Shh signaling. For instance, when MEN1 is lost, the ability to properly modulate Shh signaling can lead to the abnormal activation of downstream targets. This overactivation of Shh signaling can cause an excessive proliferation of progenitor cells in the duodenum, preventing them from differentiating into mature enteroendocrine cells [98]. Alternatively, MEN1 loss could lead to the underactivation of the Shh pathway, hindering the proper differentiation of progenitor cells into neuroendocrine cells and maintaining an undifferentiated or proliferating progenitor cell population. In addition to its role in cell differentiation, Shh signaling is involved in the spatial patterning of the gastrointestinal tract, particularly in ensuring the proper allocation of cells within the developing duodenum. The loss of MEN1 could disrupt the precise regulation of Shh signaling, leading to improper patterning of the enteroendocrine cell populations. This disruption could result in an abnormal distribution of neuroendocrine cells within the duodenum, which could further affect the secretion of critical hormones such as gastrin. Inadequate or excess gastrin production, resulting from a shift in cell fate toward an abnormal neuroendocrine phenotype, could lead to the development of gastrinomas, tumors characterized by excessive gastrin secretion. Moreover, the loss of MEN1 and the subsequent dysregulation of Shh signaling could impair the cross-talk between Shh and other signaling pathways involved in neuroendocrine differentiation, such as Notch, Wnt, and BMP. This disruption of signaling interactions can further exacerbate the imbalance in enteroendocrine cell differentiation, as these pathways typically work in concert to regulate the timing and fate of developing cells. The failure to properly integrate Shh signaling with these pathways can lead to the formation of a disorganized or dysfunctional neuroendocrine cell population, which increases the likelihood of neoplastic transformation. These cells, under conditions of disrupted differentiation and patterning, may acquire oncogenic mutations, leading to the development of neoplasms such as gastrinomas [99]. The loss of MEN1 disrupts the regulation of Shh signaling in neuroendocrine cells of the duodenum by altering its activation, cellular differentiation, and tissue patterning roles. Dysregulation of Shh signaling due to the loss of MEN1 can lead to abnormal proliferation and differentiation of progenitor cells, improper patterning of neuroendocrine cells, and increased risk of gastrinoma formation. These disruptions in the normal developmental processes of neuroendocrine cells in the duodenum highlight the critical role of MEN1 in maintaining the proper function of Shh signaling and its crucial involvement in preventing pathological conditions such as tumorigenesis [100].

### Consequences for Enteroendocrine Cells and Associated Gastrionoma Risk

The loss of MEN1, which encodes the Menin protein, significantly impacts multiple cellular pathways, including the Sonic Hedgehog (Shh) signaling pathway, which plays a vital role in regulating the differentiation and fate of neuroendocrine (enteroendocrine) cells in the duodenum. The Shh pathway is crucial for the development of these cells by maintaining the balance between progenitor cell proliferation and their differentiation into mature hormone-producing cells. When MEN1 is lost, its regulatory function over various signaling pathways, including Shh, is disrupted, leading to altered cellular processes that can have significant consequences for cell fate determination and increase the risk of tumorigenesis, such as the formation of gastrinomas. In the duodenum, Shh signaling helps to orchestrate the differentiation of progenitor cells into specialized enteroendocrine cells. MEN1 loss impairs the ability to regulate Shh signaling effectively [101]. As a result, the pathway may become overactive, causing excessive proliferation of progenitor cells and preventing their proper differentiation into mature neuroendocrine cells. On the other hand, MEN1 loss could also result in inadequate Shh pathway activation, hindering the differentiation of progenitor cells into the appropriate neuroendocrine phenotype, and maintaining an undifferentiated or proliferating population. This imbalance in Shh signaling leads to the disruption of normal cell differentiation patterns in the duodenum, contributing to a population of poorly differentiated cells that might acquire neoplastic characteristics.

Shh signaling also plays a key role in patterning the gut during development, ensuring the proper organization and allocation of neuroendocrine cells within the duodenum. Loss of MEN1 can result in abnormal Shh signaling that affects this spatial organization. As a consequence, neuroendocrine cells may be mislocalized or fail to mature into the proper cell types [102]. This disruption in cellular patterning could lead to the abnormal production of hormones, such as gastrin, by dysregulated enteroendocrine cells. Excessive or uncontrolled gastrin secretion can promote the formation of gastrinomas, a type of tumor associated with excessive gastrin production, further complicating the physiological functions of the gastrointestinal tract. Furthermore, MEN1 loss can impair the coordinated regulation between Shh signaling and other pathways that regulate neuroendocrine differentiation, such as Notch, Wnt, and BMP. These pathways interact with Shh signaling to fine-tune the differentiation process, and their disruption due to MEN1 loss can create an environment where neuroendocrine cell development is compromised. Such an environment may result in a population of cells with the potential for malignant transformation, increasing the risk of tumorigenesis. In particular, the disorganized and dysfunctional differentiation of neuroendocrine cells in the duodenum increases the likelihood that these cells may acquire mutations and develop into neoplastic cells, leading to the formation of gastrinomas. The loss of MEN1 disrupts the regulation of Shh signaling in neuroendocrine cells of the duodenum by impairing the pathway’s ability to control cell differentiation, patterning, and proliferation. This disruption results in an abnormal distribution of neuroendocrine cells, an altered cell fate, and increased production of hormones like gastrin. As a result, the loss of MEN1 creates a favorable environment for the development of gastrinomas, demonstrating the critical role of MEN1 in maintaining normal Shh signaling and preventing tumorigenesis in the gastrointestinal tract [103, 104].

### Possible Clinical Implications of Shh Pathway Dysregulation in MEN1 Patients: Enhancing Precision Diagnostics, Surveillance, and Preventive Therapeutics in Gastrinoma Care

The Sonic Hedgehog (Shh) signaling pathway plays an essential role in orchestrating the differentiation, proliferation, and spatial patterning of neuroendocrine cells in the developing and adult duodenum. In the context of MEN1 loss, this pathway becomes dysregulated due to the absence of menin, a tumor suppressor critical for maintaining transcriptional and chromatin-level control. As a result, Shh signaling may become either abnormally activated or underactive, disrupting progenitor cell dynamics and promoting conditions that lead to neuroendocrine tumorigenesis, including the development of gastrinomas. From a clinical standpoint, Shh dysregulation offers a novel and actionable biomarker for early disease detection in MEN1 mutation carriers. Since aberrant Shh signaling precedes visible tumor formation, its detection in duodenal biopsy samples via markers such as GLI1, PTCH1, and SHH ligand levels can help identify molecular mispatterning and progenitor cell proliferation before neoplasia emerges.

The use of such molecular pathology tools allows clinicians to stratify patients who may appear normal endoscopically but harbor early molecular signatures of gastrinoma risk. Moreover, the patterning role of Shh in establishing spatial organization of hormone-producing enteroendocrine cells underscores its diagnostic utility in assessing tissue architecture. In MEN1-deficient biopsies, aberrant localization of neuroendocrine cells or disrupted zonation of gastrin-secreting G cells can be subtle but detectable red flags when interpreted in conjunction with molecular profiling. Thus, integrating Shh pathway profiling into biopsy-based diagnostic workflows can significantly enhance the sensitivity and specificity of early gastrinoma detection.

In terms of surveillance, MEN1 patients with evidence of Shh dysregulation either through GLI1 overexpression, increased SHH ligand activity, or abnormal spatial organization can be identified as high-risk and followed with more frequent endoscopic and molecular monitoring. Conversely, patients with preserved Shh signaling signatures may be eligible for reduced surveillance intensity, personalizing care and minimizing unnecessary procedures.

Therapeutically, Shh pathway dysregulation presents a promising avenue for preventive intervention. Preclinical studies in other tumor types have demonstrated that Shh inhibitors (e.g., vismodegib, sonidegib) can block aberrant pathway activity and reduce tumor burden. In MEN1 patients, especially those with molecular evidence of progenitor expansion without differentiation, early Shh-targeted therapy could help re-establish normal differentiation trajectories, suppressing the expansion of pre-neoplastic cells and delaying or preventing gastrinoma onset. Furthermore, because Shh signaling interacts with Notch, Wnt, and BMP pathways, all of which are dysregulated in MEN1 loss, therapeutic strategies that combine multi-pathway modulation may yield synergistic benefits. For instance, combining Shh inhibitors with agents that restore BMP activity or modulate Notch-Wnt balance could enhance differentiation, suppress proliferation, and more effectively reduce gastrinoma risk.

Shh pathway disruption can also contribute to functional symptoms such as abnormal gastrin secretion and hyperacidity by altering the distribution and identity of gastrin-producing cells. Therefore, early detection and correction of Shh signaling abnormalities could also offer symptom control benefits in addition to cancer prevention.

## 13. MAPK/ERK

### Mutated MEN1 in Dysregulating Enteroendocrine Cells

The MAPK/ERK signaling pathway plays a crucial role in regulating cell proliferation, differentiation, and survival in various cell types, including neuroendocrine (enteroendocrine) cells in the duodenum. This pathway is activated by growth factors and extracellular signals, triggering a cascade of intracellular events that ultimately lead to the activation of ERK1/2, which then regulates gene expression associated with cell cycle progression and differentiation. In neuroendocrine cells of the duodenum, the MAPK/ERK pathway is essential for maintaining proper cellular function, differentiation, and hormonal regulation. However, the loss of MEN1, which encodes the tumor suppressor protein Menin, can disrupt this pathway and lead to various cellular abnormalities, contributing to the development of tumors such as gastrinomas. Menin, through its role in chromatin remodeling and transcriptional regulation, interacts with a number of signaling pathways, including the MAPK/ERK pathway [105]. MEN1 loss results in the loss of regulatory control over key components of this pathway, particularly affecting the expression of genes involved in the activation and modulation of ERK signaling. Under normal conditions, Menin helps modulate the activity of transcription factors such as CREB and AP-1, which are downstream of MAPK/ERK activation. When MEN1 is absent, the deregulation of these factors can lead to inappropriate activation or repression of genes involved in cell proliferation and differentiation, promoting an environment that favors tumorigenesis. The loss of MEN1 also causes disturbances in the regulation of upstream components of the MAPK/ERK pathway, such as Ras and Raf. These components are critical for initiating the signaling cascade, and their dysregulation can result in prolonged or exaggerated ERK activation. In enteroendocrine cells, this can lead to an imbalance between cellular proliferation and differentiation. Specifically, the overstimulation of the MAPK/ERK pathway can lead to excessive proliferation of precursor cells, preventing their proper differentiation into mature enteroendocrine cells. This dysregulation can result in the accumulation of undifferentiated or poorly differentiated cells, increasing the likelihood of neoplastic transformation and the development of endocrine tumors, such as gastrinomas. Furthermore, the MAPK/ERK pathway is also closely integrated with other signaling networks, such as the PI3K/Akt and JAK/STAT pathways, which also regulate cell survival, growth, and differentiation [106]. Loss of MEN1 leads to aberrant cross-talk between these pathways, amplifying the cellular responses to growth factors and cytokines. In the context of neuroendocrine cells in the duodenum, this aberrant signaling can push cells towards an undifferentiated state, thereby increasing their potential to develop into malignant tumors. This combination of dysregulated MAPK/ERK signaling and disrupted cross-talk between multiple signaling pathways creates an environment where enteric neuroendocrine cells are more prone to transformation and tumorigenesis. The loss of MEN1 in neuroendocrine cells in the duodenum results in the dysregulation of the MAPK/ERK signaling pathway, leading to altered cellular differentiation, excessive proliferation, and impaired tumor suppressor functions. This contributes to the development of neuroendocrine tumors, including gastrinomas, by promoting the accumulation of undifferentiated cells and disrupting the normal differentiation process. The inability to properly regulate the MAPK/ERK pathway and its downstream effects significantly increases the risk of tumor formation in individuals with MEN1 loss, highlighting the critical role of this signaling pathway in maintaining cellular homeostasis and preventing malignancy [107, 108].

### Consequences for Enteroendocrine Cells and Associated Gastrionoma Risk

The MAPK/ERK signaling pathway plays an essential role in regulating cell fate decisions, particularly in neuroendocrine (enteroendocrine) cells in the duodenum. This pathway governs key processes such as cell proliferation, differentiation, and survival, and is crucial for maintaining the balance between cell renewal and maturation in various tissues. In the duodenum, the MAPK/ERK pathway influences the differentiation of enteroendocrine cells, which produce various hormones that regulate digestive processes [109]. Under normal conditions, the MAPK/ERK signaling cascade is tightly regulated by multiple factors, including tumor suppressors such as MEN1. The MEN1 gene encodes the Menin protein, which plays a crucial role in maintaining cellular homeostasis, including modulating transcription factors and interacting with key signaling pathways like MAPK/ERK. When MEN1 is lost, this regulation is disrupted, leading to an imbalance in MAPK/ERK signaling. In the absence of Menin, there is an increase in the activation of MAPK/ERK components such as Ras and Raf, which results in the overactivation of ERK signaling. This unchecked signaling can alter the normal cell fate decisions in neuroendocrine cells, specifically affecting their differentiation into mature enteroendocrine cells. Instead of undergoing normal differentiation, precursor cells may undergo excessive proliferation, pushing them toward a more undifferentiated or hyperproliferative state. This leads to a failure in the proper development of enteroendocrine cell populations, which can promote the accumulation of precursor or immature cells within the duodenal epithelium. Additionally, the sustained activation of the MAPK/ERK pathway due to MEN1 loss can lead to cellular transformation and malignant progression. In neuroendocrine cells, this dysregulation may contribute to the development of tumors, such as gastrinomas, which are commonly associated with MEN1 mutations. Gastrinomas arise from enteroendocrine cells that fail to properly differentiate and instead proliferate uncontrollably. The abnormal activation of MAPK/ERK signaling caused by MEN1 loss can exacerbate this process by driving continued cell cycle progression and blocking normal differentiation cues. As a result, undifferentiated or poorly differentiated cells accumulate, and the potential for neoplastic transformation increases [110]. Furthermore, MEN1 loss-induced dysregulation of the MAPK/ERK pathway can have broader effects by disrupting the normal interplay between MAPK/ERK and other signaling pathways, such as PI3K/Akt and Notch, that regulate cell survival and differentiation. This cross-talk between pathways may amplify the proliferative signals or further inhibit differentiation, further tipping the balance toward tumorigenesis. As the MAPK/ERK pathway interacts with various growth factors and cytokine signaling pathways, the loss of MEN1 impairs the feedback mechanisms that normally keep cell growth and differentiation in check, leading to an increased risk of tumorigenesis, particularly in the context of gastrinomas. The loss of MEN1 disrupts the regulation of the MAPK/ERK signaling pathway, resulting in aberrant cell fate determination in neuroendocrine cells in the duodenum. This disruption leads to increased proliferation and decreased differentiation, creating an environment that is prone to the development of gastrinomas. By driving abnormal cell growth and impairing normal differentiation processes, MEN1 loss enhances the likelihood of neoplastic transformation in these cells, significantly increasing the risk of gastrinoma formation in affected individuals [111, 112].

### Possible Clinical Implications of MAPK/ERK Pathway Dysregulation in MEN1-Associated Gastrinoma: Toward Early Detection, Precision Surveillance, and Targeted Therapy

The MAPK/ERK signaling pathway is central to regulating the balance between proliferation and differentiation in duodenal neuroendocrine cells. In healthy conditions, this pathway is tightly modulated by growth factors and transcriptional regulators, including menin, the tumor suppressor encoded by the MEN1 gene. Loss of MEN1 disrupts this regulation, leading to chronic ERK pathway activation, impaired differentiation, and uncontrolled precursor cell expansion hallmarks of early tumorigenesis in gastrinoma development. From a diagnostic and clinical surveillance standpoint, MEN1-associated MAPK/ERK dysregulation provides a valuable molecular biomarker for early detection of pre-neoplastic changes in MEN1 patients. Biomarkers such as phospho-ERK1/2, elevated expression of immediate early genes (e.g., c-FOS, c-JUN), and nuclear localization of ELK1 can be evaluated in duodenal biopsy specimens.

These markers reflect pathway hyperactivation and can signal the presence of proliferative but undifferentiated neuroendocrine cells well before visible tumors arise. Importantly, identifying MAPK/ERK activation in biopsy samples enables patient stratification for surveillance intensity. MEN1 patients showing high phospho-ERK or overexpression of ERK targets may benefit from more frequent endoscopic and molecular monitoring, as they are at higher risk for gastrin-secreting tumorigenesis. Conversely, those without detectable ERK dysregulation may be suitable for standard-risk surveillance pathways.

Therapeutically, MAPK/ERK dysregulation is a clinically actionable pathway. Several inhibitors targeting MEK or ERK (e.g., trametinib, cobimetinib) are FDA-approved in other cancers and may have preventive or disease-modifying potential in high-risk MEN1 patients, particularly those with early biomarker elevation but no radiological tumor burden. Such chemopreventive strategies could suppress proliferative signaling and support re-engagement of differentiation programs in neuroendocrine progenitors. Moreover, the MAPK/ERK pathway does not function in isolation. It interacts extensively with PI3K/Akt, JAK/STAT, and Notch all of which are influenced by MEN1 loss. This suggests that combination therapies or multi-pathway modulators may more effectively correct the imbalance in cell fate determination, especially in cases where MAPK/ERK activation is synergized by co-dysregulation of other oncogenic axes. In addition to tumor prevention, modulating the MAPK/ERK pathway may have functional benefits. As this pathway also governs hormonal outputs and differentiation of hormone-producing cells, restoring normal MAPK activity could reduce aberrant gastrin secretion, mitigating symptoms of Zollinger-Ellison syndrome and improving gastrointestinal function in MEN1 patients even before tumors manifest.

## 14. The mTOR Pathway

### Mutated MEN1 in Dysregulating Enteroendocrine Cells

The mTOR signaling pathway is a critical regulator of cell growth, proliferation, and differentiation, playing a key role in controlling cellular responses to nutrients, growth factors, and stress signals. In neuroendocrine (enteroendocrine) cells in the duodenum, mTOR regulates various cellular processes, including protein synthesis, cell metabolism, and the maturation of neuroendocrine cells [113]. The pathway is tightly controlled by upstream tumor suppressors like MEN1, whose encoded protein Menin interacts with multiple signaling pathways, including mTOR, to maintain cellular homeostasis and prevent uncontrolled growth. Loss of MEN1 disrupts the regulation of mTOR signaling, leading to an imbalance in its activation. Menin functions as a negative regulator of mTOR in certain contexts, and its loss removes this checkpoint, resulting in the hyperactivation of mTORC1, a key complex within the mTOR pathway that regulates cell growth and metabolism. The dysregulation of mTORC1 activity due to MEN1 loss leads to an increase in protein synthesis and cellular growth, while inhibiting autophagy and other processes that typically counteract excessive growth [114]. In neuroendocrine cells, this disruption in mTOR signaling impairs the normal differentiation process by promoting cell cycle progression and inhibiting differentiation cues, leading to the accumulation of immature or undifferentiated cells. In the duodenum, the loss of MEN1-induced dysregulation of mTOR signaling may push enteroendocrine precursor cells toward uncontrolled proliferation, as mTORC1 activation also enhances the expression of genes involved in cell cycle progression. Additionally, mTORC1 signaling promotes the upregulation of anabolic processes, including the synthesis of lipids and proteins, further supporting cellular growth at the expense of proper differentiation. These changes in cellular behavior can result in the aberrant expansion of proliferating cells, thereby contributing to an increased risk of tumorigenesis in the duodenal neuroendocrine cell population. This is particularly significant in the context of MEN1-related conditions, where tumorigenesis is frequently observed in neuroendocrine tissues, including the development of gastrinomas. The hyperactivation of mTOR signaling also interacts with other pathways that are critical for cell fate determination, such as the PI3K/Akt and MAPK/ERK pathways, potentially exacerbating the proliferative effects seen in MEN1 loss. The cross-talk between mTOR and these pathways may further enhance cell survival and proliferation while impairing normal differentiation signals. In addition, the dysregulation of mTOR signaling may affect the responsiveness of neuroendocrine cells to external cues, including hormones and growth factors, altering their ability to function properly in the duodenum.

Furthermore, mTORC1 signaling is known to inhibit autophagy, a process that is crucial for maintaining cellular homeostasis by degrading damaged or unnecessary components. The inhibition of autophagy, resulting from MEN1 loss and mTOR activation, can lead to the accumulation of dysfunctional cellular components and disrupt the balance of cellular turnover. In neuroendocrine cells, this further impairs differentiation and promotes the development of abnormal cell populations that are prone to neoplastic transformation. The loss of MEN1 leads to the dysregulation of mTOR signaling, resulting in the hyperactivation of mTORC1 and disruption of normal neuroendocrine cell differentiation in the duodenum [115]. This dysregulation favors uncontrolled proliferation, inhibits proper cell maturation, and increases the likelihood of tumorigenesis, particularly in the form of gastrinomas. By promoting abnormal cell growth and inhibiting essential differentiation and homeostatic mechanisms, MEN1 loss creates an environment conducive to the development of neuroendocrine tumors in the duodenum [116].

### Consequences for Enteroendocrine Cells and Associated Gastrionoma Risk

The mTOR (mechanistic target of rapamycin) pathway plays a crucial role in regulating cellular growth, metabolism, and differentiation, and is particularly significant in the development and function of neuroendocrine cells, including enteroendocrine cells in the duodenum. mTOR functions as a central hub in integrating various signals from growth factors, nutrients, and cellular stress, directing cellular responses through its two main complexes, mTORC1 and mTORC2. The activity of mTORC1, in particular, is tightly regulated by tumor suppressors, including MEN1, which encodes the Menin protein. Menin interacts with various signaling pathways, including mTOR, to prevent uncontrolled cell growth and ensure proper cellular differentiation [117]. Therefore, the loss of MEN1 can have profound effects on mTOR signaling, disrupting cellular processes that are vital for maintaining normal enteroendocrine cell function. When MEN1 is lost, the regulation of mTOR signaling is disrupted, particularly mTORC1, which becomes hyperactivated. Menin normally functions to restrict mTORC1 activation, and its loss removes this inhibitory control. As a result, mTORC1 signaling is upregulated, leading to increased protein synthesis, cell cycle progression, and cell growth. These changes are particularly significant in neuroendocrine cells of the duodenum, where mTORC1 activation can promote the expansion of immature or proliferating cells, rather than supporting proper differentiation. This disruption of the normal differentiation process shifts the fate of these cells toward proliferation, creating an environment conducive to tumorigenesis. Hyperactivation of mTORC1 also inhibits autophagy, a process that plays a crucial role in cellular homeostasis by eliminating damaged proteins and organelles. The failure to clear cellular debris due to inhibited autophagy can further contribute to the accumulation of dysfunctional components, worsening the overall cellular environment and potentially contributing to neoplastic transformation. In the context of enteroendocrine cells in the duodenum, the loss of MEN1 and the subsequent mTORC1 hyperactivation disrupt normal cell fate decisions by promoting proliferation at the expense of differentiation [118]. The increased mTOR activity accelerates cell cycle progression and upregulates metabolic processes that support cell growth, while simultaneously inhibiting processes that would normally lead to differentiation and functional specialization of enteroendocrine cells. This imbalance is particularly problematic in tissues such as the duodenum, where specialized neuroendocrine cells are critical for hormone secretion and digestion. Inappropriate expansion of progenitor-like cells that fail to mature properly can lead to the accumulation of undifferentiated or functionally impaired cells, which can then progress toward neoplastic states. The hyperactivation of mTORC1 also has a significant effect on other signaling pathways that regulate cellular fate. These include the PI3K/Akt and MAPK/ERK pathways, both of which promote cell survival, growth, and proliferation. The cross-talk between mTORC1 and these pathways further amplifies the proliferative signals, making the affected cells more resistant to differentiation cues. As a result, the loss of MEN1 and the consequent mTOR dysregulation tip the balance of cellular fate toward unregulated proliferation, increasing the risk of tumor formation. Moreover, mTORC1 signaling also affects cellular responses to external cues, such as hormones and growth factors, that are crucial for normal enteroendocrine cell function. In neuroendocrine cells, the loss of MEN1-mediated regulation of mTOR could impair the cells’ ability to respond appropriately to these signals, contributing to abnormal cell behaviors, including the dysregulated secretion of hormones such as gastrin. The increased production of gastrin, often associated with hyperplasia and tumor formation in the duodenum, is a hallmark feature of gastrinoma, a type of neuroendocrine tumor. Therefore, the loss of MEN1 and the resulting mTORC1 dysregulation create an environment that promotes the expansion of proliferating cells while impairing their differentiation and function, setting the stage for the development of gastrinomas. This disrupts the regulation of the mTOR pathway, particularly mTORC1, leading to increased cell proliferation and reduced differentiation in duodenal enteroendocrine cells. This dysregulation creates a cellular environment that favors the development of tumors, particularly gastrinomas, by promoting abnormal cell growth and inhibiting differentiation processes. The loss of MEN1 thus increases the risk of neuroendocrine tumors in the duodenum by driving the expansion of proliferative, undifferentiated cell populations that are prone to neoplastic transformation [119, 120].

### Possible Clinical Implications of mTOR Pathway Dysregulation in MEN1-Associated Gastrinoma: From Molecular Insight to Patient-Centered Interventions

The mTOR signaling pathway particularly through its mTORC1 complex is a master regulator of cell growth, proliferation, metabolism, and differentiation, all of which are crucial for maintaining homeostasis in duodenal enteroendocrine cells. In the setting of MEN1 gene loss, this pathway becomes hyperactivated, disrupting the balance between proliferation and differentiation, and thereby promoting a pro-tumorigenic state. These mechanistic insights have direct clinical relevance and can be translated into meaningful advances in diagnostic precision, surveillance strategies, and therapeutic interventions for MEN1 patients at risk of gastrinoma.

The hyperactivation of mTORC1 in MEN1-deficient tissues can be identified through immunohistochemical detection of phosphorylated S6 kinase (pS6K) or 4E-BP1, which serve as surrogate markers for mTOR activity. Inclusion of these markers in routine biopsy panels from MEN1 patients especially during endoscopic surveillance can allow early identification of molecular lesions prior to visible tumor formation. This approach enables preclinical risk detection, supporting the early stratification of patients into high-risk surveillance cohorts.

MEN1 patients exhibiting elevated mTOR activity in duodenal mucosal biopsies can be classified as high-risk for gastrinoma development. This stratification allows for tailored surveillance frequencies, such as annual endoscopy or molecular monitoring, rather than a one-size-fits-all approach. Patients without significant mTOR activation may be eligible for less intensive follow-up, reducing healthcare burden and patient anxiety.

Hyperactive mTOR signaling is clinically actionable. mTOR inhibitors such as everolimus and temsirolimus, already in use for pancreatic neuroendocrine tumors (pNETs), represent promising candidates for chemopreventive or early interventional use in MEN1 patients showing early mTOR-driven molecular alterations. By suppressing mTORC1 activity, these agents may promote differentiation, reduce proliferation, and delay or prevent tumor initiation. This is particularly critical in patients with non-resectable microscopic gastrinomas or those undergoing conservative surveillance.

Beyond tumor prevention, mTOR pathway correction may restore differentiation in neuroendocrine precursor cells, thereby improving the functional integrity of enteroendocrine cell populations. This could reduce excessive gastrin secretion seen in early gastrinoma pathogenesis, offering not just oncologic but also symptom-control benefits, a significant quality-of-life consideration in MEN1 patients with Zollinger-Ellison syndrome. Patients showing co-activation of these pathways may benefit from dual or combination therapies (e.g., mTOR + MEK inhibitors), which could more robustly restore cellular balance than monotherapy. This multi-target approach can be tailored based on biomarker panels from duodenal biopsies, paving the way for personalized, pathway-informed management plans. The dysregulation of the mTOR pathway following MEN1 loss represents both a central mechanism in gastrinoma development and a clinically targetable vulnerability. By incorporating mTOR-based diagnostics, surveillance markers, and targeted therapies, clinicians can advance precision care for MEN1 patients, potentially intercepting tumorigenesis before it becomes clinically manifest, and improving both outcomes and quality of life.

## Discussion

### Possible Advances in Future Patient Care Through This Study

1. **Improved Biopsy-Based Diagnostic Approaches**

a. Molecular Marker Identification: This study systematically identifies a set of transcription factors (e.g., NEUROG3, ASCL1, PAX4, PAX6, ISL1, NKX2.2, INSM1) and signaling pathways (Notch, Wnt, BMP, Shh, MAPK/ERK, mTOR) that are dysregulated in the duodenum following MEN1 loss. These markers are measurable in biopsy samples and can serve as early indicators of molecular transformation.
b. Immunohistochemical (IHC) Utility: Several markers (e.g., phospho-S6K, pERK1/2, INSM1, ASCL1) have practical IHC potential and can be used to assess pre-neoplastic changes in MEN1 patients during routine endoscopic biopsies.
2. **Enhanced Screening and Early Detection**

Preclinical Biomarker Surveillance: The study highlights how MEN1 loss leads to dysregulation before tumor formation, offering a molecular window for early screening using gene expression profiling or IHC.
Cell Lineage Tracing Insight: By focusing on transcriptional regulators and their perturbations, this study enhances understanding of which enteroendocrine cell subtypes are more vulnerable (e.g., G-cell lineage), helping prioritize targets for screening.
3. **Personalized Surveillance Strategies**

Risk Stratification Based on Pathway Activation: Patients showing activation of high-risk pathways (e.g., mTORC1, MAPK/ERK, Notch suppression) can be stratified for more intensive surveillance.
Guidance for Surveillance Intervals: Insights from this study support tailored surveillance schedules based on molecular risk, potentially improving early intervention while reducing unnecessary procedures in low-risk individuals.
4. **New Potential Therapeutic Targets**

Targetable Signaling Pathways: The study identifies multiple clinically actionable pathways such as mTORC1, MAPK/ERK, and Notch that could be therapeutically modulated to prevent or delay gastrinoma formation.
Rationale for Chemoprevention: The findings provide a mechanistic rationale for early use of mTOR inhibitors or pathway modulators in molecularly high-risk MEN1 patients, even before radiologic tumor detection.
5. **Restoration of Cell Differentiation and Function**

i. Therapeutic Differentiation Goals: By identifying the transcriptional misregulation that keeps progenitor cells from fully differentiating, the study opens the possibility of differentiation-restoring therapies, which may limit neoplastic transformation and restore hormonal balance.
ii. Reduction of Gastrin Hypersecretion: Improved understanding of upstream regulators of gastrin-producing G cells offers the potential to mitigate Zollinger-Ellison syndrome symptoms through earlier or better-targeted treatments.
6. **Foundation for Precision Medicine in MEN1**

Tissue-Specific Molecular Profiling: The detailed framework of MEN1’s molecular effects in the duodenum lays the groundwork for individualized monitoring based on patient-specific expression profiles.
Future Biomarker Panels: This study provides a platform for developing combinatorial biomarker panels for clinical translation—blending transcription factor expression, pathway activation, and histologic changes.

The findings of this research have significant clinical implications for the diagnosis, management, and treatment of gastrinomas, particularly those linked to the loss of MEN1. Understanding how MEN1 loss disrupts the developmental regulators of neuroendocrine (enteroendocrine) cells in the duodenum provides a more detailed mechanistic framework for the early detection of gastrinomas, allowing for more precise diagnostic tools and biomarkers. By identifying the key transcription factors and signaling pathways that are altered in MEN1-deficient neuroendocrine cells, targeted therapies could be developed to restore proper cellular differentiation and proliferation patterns, potentially preventing the formation of gastrinomas or reducing tumor burden. Clinically, this research also opens avenues for personalized treatments based on the specific molecular signatures of MEN1 loss in neuroendocrine tumors. For example, therapies that target the dysregulated MAPK/ERK or mTOR pathways might help reverse some of the tumorigenic effects associated with MEN1 loss. Moreover, the exploration of small molecules or gene therapies that can specifically correct the dysfunction in developmental regulators like NEUROG3, ASCL1, or PAX4 could provide new treatment strategies to prevent or manage gastrinoma development in patients with MEN1 mutations. In terms of future research directions, the focus should shift towards refining our understanding of how each developmental regulator individually and synergistically contributes to the progression of gastrinomas. Longitudinal studies exploring the early stages of neuroendocrine cell development in MEN1-deficient models could reveal potential intervention points before tumor formation occurs. Furthermore, investigating the interactions between MEN1 and other genes involved in endocrine signaling will offer deeper insights into the complex regulatory networks governing neuroendocrine cell fate. Finally, clinical trials assessing the efficacy of therapies targeting the specific pathways identified in this study will be essential for validating these approaches and advancing the treatment of MEN1-associated neuroendocrine tumors.

The MEN1 mutation leads to a dysregulation of both proliferation and differentiation processes in neuroendocrine (enteroendocrine) cells within the duodenum, contributing to the development of gastrinoma. MEN1 is a key tumor suppressor gene, and its loss disrupts the delicate balance between the proliferation and differentiation of these cells by altering various developmental regulators and signaling pathways. In normal cellular processes, neuroendocrine cells undergo a tightly regulated process of differentiation, driven by transcription factors such as NEUROG3, ASCL1, PAX4, PAX6, ISL1, NKX2.2, INSM1, and ARX, which guide the development of specialized enteroendocrine cells with distinct functions, including hormone production. However, when MEN1 is lost, these regulatory pathways are disrupted, leading to the loss of differentiation cues while simultaneously enhancing cellular proliferation. For instance, in the absence of MEN1, NEUROG3, a crucial transcription factor for enteroendocrine cell differentiation, may fail to activate the necessary downstream programs for proper differentiation. Instead of maturing into functional enteroendocrine cells, the progenitors may proliferate uncontrollably, driven by other dysregulated factors. Similarly, transcription factors like ASCL1 and PAX6, which are pivotal for the establishment of neuroendocrine lineages, are likely to be improperly regulated in MEN1-deficient cells, further skewing the differentiation process. The loss of MEN1 may thus increase the proportion of undifferentiated or less differentiated cells that retain the capacity for rapid proliferation. In parallel, key signaling pathways such as Notch, Wnt, BMP, Shh, MAPK/ERK, and mTOR, which are involved in regulating cell fate decisions, become altered due to MEN1 mutation. Notch signaling, for example, plays a significant role in the maintenance of stemness and the regulation of differentiation in various tissues, including the gut. In the absence of MEN1, Notch signaling may be dysregulated, promoting excessive self-renewal and limiting differentiation, thereby increasing the risk of hyperplastic cell growth. Likewise, Wnt and BMP signaling pathways, both of which influence cellular differentiation, are likely disrupted in MEN1-deficient cells, contributing to uncontrolled cell proliferation. The MAPK/ERK and mTOR pathways, both central to cell growth, metabolism, and survival, are often hyperactivated in the context of MEN1 loss. In MEN1-deficient neuroendocrine cells, this hyperactivation leads to enhanced proliferation rather than differentiation, further promoting the formation of tumors such as gastrinomas. The dysregulation of these pathways can result in the accumulation of progenitor-like cells that fail to fully differentiate into their mature, hormone-producing forms, creating an environment ripe for tumorigenesis. Ultimately, the loss of MEN1 alters a network of developmental regulators and signaling pathways in neuroendocrine cells of the duodenum, driving a shift toward uncontrolled proliferation rather than the appropriate differentiation of enteroendocrine cells. This disruption results in the accumulation of undifferentiated, proliferative cells, increasing the risk of forming gastrinomas. The development of these tumors is likely driven by the inability to properly regulate differentiation, alongside unchecked proliferation fueled by dysregulated signaling pathways.

### Key Findings

1. MEN1 Loss Alters Neuroendocrine Cell Fate Decisions: MEN1 deficiency dysregulates developmental transcription factors and signaling pathways, shifting the balance from normal differentiation toward aberrant proliferation in duodenal enteroendocrine cells.
2. Dysregulation of Early Cell Fate Specifiers: NEUROG3 and ASCL1, key regulators of enteroendocrine lineage commitment, are overexpressed or sustained abnormally under MEN1 loss, leading to expansion of immature neuroendocrine progenitors.
3. Disruption of Differentiation Pathways: PAX4, PAX6, ISL1, and NKX2.2 show altered expression patterns, impairing terminal differentiation of neuroendocrine cells and favoring a progenitor-like or hyperplastic state conducive to tumorigenesis.
4. Aberrant Activation of Lineage Drivers: INSM1 and ARX, involved in terminal differentiation and subtype specification, are dysregulated, promoting instability in cell identity and potential trans-differentiation toward gastrin-secreting phenotypes.
5. Signaling Pathway Dysregulation: Loss of MEN1 leads to misregulation of major pathways: Notch: suppression favors neuroendocrine differentiation but unchecked expansion. Wnt, BMP, Shh: altered gradients affect progenitor maintenance and differentiation.
6. MAPK/ERK and mTOR: upregulation supports hyperproliferation and survival of tumor-prone EECs.
7. Shift Toward Proliferation Over Differentiation: MEN1 mutation promotes mitogenic signaling and impairs cell cycle exit, driving excessive EEC proliferation without proper maturation.
8. Increased Risk of Gastrinoma Formation: The cumulative dysregulation of these regulators and pathways fosters an environment that favors gastrin-secreting cell expansion and neoplastic transformation, ultimately predisposing to gastrinoma.

### Future Directions

Building on the findings of this study, future research should focus on the validation of the identified transcription factors and signaling pathways (e.g., NEUROG3, ASCL1, PAX4/6, mTOR, MAPK/ERK, Notch) as potential biomarkers for early gastrinoma risk stratification in MEN1 patients. This could involve biopsy-based molecular profiling of duodenal tissue from MEN1 carriers before tumor onset. Additionally, prospective cohort studies and functional genomic experiments are warranted to determine the causative role and timing of these molecular disruptions in tumor initiation. There is also a strong rationale for exploring targeted therapeutics that modulate these dysregulated pathways to delay or prevent tumor development. Integrating these markers into endoscopic surveillance protocols may enable a shift toward precision surveillance strategies, ultimately improving outcomes through early intervention in high-risk MEN1 populations.

## Conclusion

This study provides critical clinical insight into how MEN1 loss disrupts key transcription factors and signaling pathways that regulate duodenal enteroendocrine cell differentiation, predisposing patients to gastrinoma development. By identifying molecular drivers such as NEUROG3, ASCL1, and mTOR, the study offers a foundation for biopsy-based early detection and risk stratification in MEN1 patients. These findings can inform targeted surveillance protocols and pave the way for therapeutic interventions aimed at restoring normal differentiation. The integration of these molecular insights into clinical practice holds promise for earlier diagnosis, improved patient monitoring, and potentially reduced gastrinoma-related complications.

The loss of MEN1 has a profound impact on the developmental regulators of neuroendocrine (enteroendocrine) cells in the duodenum, leading to the dysregulation of key signaling pathways and transcription factors (TFs) involved in the differentiation, growth, and function of these cells. MEN1, through its encoded protein Menin, plays a crucial role in regulating several developmental pathways and ensuring proper cellular differentiation and homeostasis. When MEN1 is lost, its regulatory functions are impaired, disrupting the balance between cell proliferation and differentiation, which is crucial for maintaining the normal function of enteroendocrine cells. This loss leads to the hyperactivation of multiple pathways, promoting aberrant cell proliferation, altered differentiation, and the eventual formation of tumors, such as gastrinomas. Transcription factors such as NEUROG3, ASCL1, PAX4, PAX6, ISL1, NKX2.2, INSM1, and ARX are key regulators of neuroendocrine cell fate and differentiation in the duodenum. MEN1 loss impairs their normal expression and activity, disrupting the fine balance required for proper neuroendocrine differentiation. NEUROG3, for instance, is a critical regulator of pancreatic and enteroendocrine cell development, and its dysregulation in the absence of MEN1 can lead to defective enteroendocrine cell differentiation. Similarly, ASCL1 and PAX4 are essential for the development and maintenance of endocrine cells, and their dysregulated expression due to MEN1 loss can result in a failure of proper cell fate decisions, driving the expansion of progenitor-like cells that are prone to neoplastic transformation. PAX6 and ISL1 also play important roles in controlling the differentiation of neuroendocrine cells in the duodenum, and their loss of regulation can promote the expansion of undifferentiated cells, increasing the risk of tumorigenesis. The Notch, Wnt, BMP, Shh, MAPK/ERK, and mTOR signaling pathways, all of which are heavily influenced by MEN1, are integral in controlling neuroendocrine cell differentiation and function. MEN1’s loss leads to the hyperactivation of several of these pathways, including mTOR, MAPK/ERK, and Wnt, resulting in excessive cell proliferation and impaired differentiation. mTORC1 hyperactivation, in particular, drives an imbalance between cell growth and differentiation, inhibiting processes like autophagy that are critical for maintaining cellular integrity. The Wnt and Notch pathways, essential for regulating stem cell proliferation and differentiation, are also disrupted, further tipping the balance toward abnormal cell proliferation. These changes in signaling disrupt the normal fate determination of neuroendocrine cells, leading to the expansion of proliferative, undifferentiated cell populations that are prone to neoplastic transformation. Similarly, the BMP and Shh pathways, both of which help control cell differentiation and fate decisions, are compromised, further contributing to the failure of proper enteroendocrine cell differentiation. The dysregulation of these pathways and transcription factors due to the loss of MEN1 ultimately results in the development of gastrinoma. Gastrinomas are neuroendocrine tumors characterized by the overproduction of gastrin, a hormone involved in regulating gastric acid secretion. The loss of MEN1, by disrupting the regulatory networks controlling the fate of neuroendocrine cells, leads to the expansion of precursor cells that fail to differentiate into specialized enteroendocrine cells. This aberrant expansion results in an overproduction of gastrin and the formation of gastrinomas, which are often associated with MEN1 syndrome. The combined effects of disrupted cell differentiation, uncontrolled proliferation, and impaired signaling regulation create an environment that favors the development of these neuroendocrine tumors, highlighting the critical role of MEN1 in maintaining the balance between cell differentiation and proliferation in the duodenum. The loss of MEN1 disrupts a complex network of transcription factors and signaling pathways crucial for the normal development and differentiation of neuroendocrine (enteroendocrine) cells in the duodenum. This disruption leads to a failure in the proper regulation of cell fate decisions, resulting in the expansion of undifferentiated, proliferating cells and ultimately increasing the risk of gastrinoma formation. MEN1’s regulatory role is thus essential for maintaining the delicate balance required for normal cell differentiation, and its loss contributes significantly to the pathogenesis of gastrinoma in the context of neuroendocrine cell development. Ultimately, this study advances precision medicine approaches in the management of MEN1-associated neuroendocrine tumors.

## Abbreviations

MEN1: Multiple Endocrine Neoplasia Type 1
NEUROG3: Neurogenin 3
ASCL1: Achaete-Scute Family
BHLH: Transcription Factor 1
PAX4: Paired Box Gene 4
PAX6: Paired Box Gene 6
ISL1: ISL LIM Homeobox 1
NKX2.2: NK2 Homeobox 2
INSM1: Insulinoma-Associated Protein 1
ARX: Aristaless Related Homeobox
BMP: Bone Morphogenetic Protein
Shh: Sonic Hedgehog
MAPK/ERK: Mitogen-Activated Protein Kinase / Extracellular Signal-Regulated Kinase
mTOR: Mechanistic Target of Rapamycin
TF: Transcription Factor
EECs: Enteroendocrine Cells
NECs: Neuroendocrine Cells

## Declarations

### Ethics declarations

#### Ethics approval and consent to participate

Not applicable.

### Consent for publication

Not applicable.

### Data Availability statement

All data generated or analyzed during this study are included in this article.

### Competing interests

The authors declare that they have no competing interests.

## Funding

I declare that there was not any source of funding for this research work.

### Acknowledgements

We are thankful to Muhammad Danial Yaqub (MDY), MBBS, for coordinating this research project among multiple co-authors.

## Authors’ Information

**1. Ovais Shafi (OS)\*** is the author of the study and was involved in the idea, concept, design, and methodology of the study, literature search and references. He did the writing, editing, and revision of the manuscript. He was involved in drawing the findings, results, conclusions, implications of the study, interpretation of the data and was involved in all aspects of the study. He prepared and wrote discussion, results, conclusions and all areas of the study. OS extracted and analyzed the data. He was involved in critical evaluation, audit of every aspect of the study, data extraction, adherence of the study to relevant PRISMA guidelines, limitations of the study, references, and all others. He was involved in drawing PRISMA Flow Diagram. The author read and approved the manuscript.

He investigated how the loss of MEN1 on developmental regulators (NEUROG3, ASCL1, PAX4, PAX6, ISL1, NKX2.2, INSM1, ARX, Notch, Wnt, BMP, Shh, MAPK/ERK, mTOR) of duodenal enteroendocrine cells, results in gastrinoma development. He contributed towards all the investigated areas.

**Ovais Shafi (OS)*,** MBBS - Sindh Medical College - Dow University of Health Sciences, Karachi, Pakistan. He aspires to become an eminent ‘Physician Scientist’. He is devoted to the research in disease development mechanisms, disease origins and therapeutics. OS is also passionate about multiple research areas including clinical trials, clinical medicine, therapeutics, regenerative medicine, precision medicine including gene therapies, finding disease specific targets for gene therapy, role of disease genomics and epigenetics in diagnosis, management, and therapeutics development. He is dedicated to the field of research and clinical medicine.

**Corresponding author: OS**

Correspondence to Ovais Shafi

**2. Aakash (AA)** is the co-author of the study. He contributed to the results and conclusions of the study, also contributed to the writing and editing of these sections. He also contributed to the references. He contributed to investigating how the loss of MEN1 on developmental regulators (NEUROG3, ASCL1, PAX4, PAX6, ISL1, NKX2.2, INSM1, ARX, Notch, Wnt, BMP, Shh, MAPK/ERK, mTOR) of duodenal enteroendocrine cells, results in gastrinoma development. He contributed towards all the investigated areas.

Aakash MD, currently working in Florida State University Cape Coral Hospital and he is MBBS from Sindh Medical College - Dow University of Health Sciences, Karachi, Pakistan. He is passionate about pursuing fellowship in gastroenterology. His goal is also to translate emerging findings in research of disease development mechanisms/ origins into their clinical implications.

**3. Luqman Naseer Virk (LNV)** is the co-author of the study. He contributed to the results and conclusions of the study, also contributed to the writing and editing of these sections. He also contributed to the references. He contributed to investigating how the loss of MEN1 on developmental regulators (NEUROG3, ASCL1, PAX4, PAX6, ISL1, NKX2.2, INSM1, ARX, Notch, Wnt, BMP, Shh, MAPK/ERK, mTOR) of duodenal enteroendocrine cells, results in gastrinoma development. He contributed towards all the investigated areas.

Luqman Naseer Virk (LNV), MBBS - Sindh Medical College - Dow University of Health Sciences, Karachi, Pakistan. He is passionate doctor and is enthusiastic about research in medicine/surgery and disease development mechanisms including gastroenterology, cardiology, neurodegenerative diseases, oncogenesis and others.

**4. Mustafa Ali Hamid (MAH)** is the co-author of the study. He contributed to the results and conclusions of the study, also contributed to the writing and editing of these sections. He also contributed to the references. He contributed to investigating how the loss of MEN1 on developmental regulators (NEUROG3, ASCL1, PAX4, PAX6, ISL1, NKX2.2, INSM1, ARX, Notch, Wnt, BMP, Shh, MAPK/ERK, mTOR) of duodenal enteroendocrine cells, results in gastrinoma development. He contributed towards all the investigated areas.

He is currently working at Lincoln County Hospital, Lincoln, United Kingdom.

**5. Sohaib Khalid (SK)** is the co-author of the study. He contributed to the results and conclusions of the study, also contributed to the writing and editing of these sections. He also contributed to the references. He contributed to investigating how the loss of MEN1 on developmental regulators (NEUROG3, ASCL1, PAX4,

PAX6, ISL1, NKX2.2, INSM1, ARX, Notch, Wnt, BMP, Shh, MAPK/ERK, mTOR) of duodenal enteroendocrine cells, results in gastrinoma development. He contributed towards all the investigated areas.

He is currently working at Lincoln County Hospital, Lincoln, United Kingdom.

**6. Deepak Kumar (DK)** is the co-author of the study. He contributed to the results and conclusions of the study, also contributed to the writing and editing of these sections. He also contributed to the references. He contributed to investigating how the loss of MEN1 on developmental regulators of duodenal enteroendocrine cells, results in gastrinoma development. He contributed towards 11 out of 14 investigated areas in relation to the study: NEUROG3, ASCL1, PAX4, PAX6, ISL1, NKX2.2, INSM1, ARX, Notch, Wnt, BMP. He is currently working at Zucker School of Medicine at Hofstra, Northwell at Mather Hospital, NY, USA.

**7. Raveena (RA)** is the co-author of the study. She contributed to the results and conclusions of the study, also contributed to the writing and editing of these sections. She also contributed to the references. She contributed to investigating how the loss of MEN1 on developmental regulators (NEUROG3, ASCL1, PAX4, PAX6, ISL1, NKX2.2, INSM1, ARX, Notch, Wnt, BMP, Shh, MAPK/ERK, mTOR) of duodenal enteroendocrine cells, results in gastrinoma development. She contributed towards all the investigated areas.

Raveena, MBBS - Sindh Medical College – Jinnah Sindh Medical University, Karachi, Pakistan. She is passionate about research in surgery and disease development mechanisms including neurodegenerative diseases, oncogenesis and others. She is ECFMG Certified. She is passionate about residency in Internal Medicine/Surgery. Her goal is to make significant impact in the field of Research.

**8. Deepika Kumari Kataria (DKK)** is the co-author of the study. She contributed to the results and conclusions of the study, also contributed to the writing and editing of these sections. She also contributed to the references. She contributed to investigating how the loss of MEN1 on developmental regulators of duodenal enteroendocrine cells, results in gastrinoma development. She contributed towards 7 out of 14 investigated areas in relation to the study: NEUROG3, ASCL1, PAX4, PAX6, ISL1, NKX2.2, INSM1.

She is MBBS from Chandka Medical College SMBBMU Larkana, Sindh, Pakistan.

**9. Muhammad Danial Yaqub (MDY)** is the co-author of the study. He contributed to the results and conclusions of the study, also contributed to the writing and editing of these sections. He also contributed to the references. He contributed to investigating how the loss of MEN1 on developmental regulators (NEUROG3, ASCL1, PAX4, PAX6, ISL1, NKX2.2, INSM1, ARX, Notch, Wnt, BMP, Shh, MAPK/ERK, mTOR) of duodenal enteroendocrine cells, results in gastrinoma development. He contributed towards all the investigated areas.

Muhammad Danial Yaqub is currently working at Lincoln County Hospital, Lincoln, United Kingdom.

**10. Rahimeen Rajpar (RR)** is the co-author of the study. She contributed to the results and conclusions of the study, also contributed to the writing and editing of these sections. She also contributed to the references. She contributed to investigating how the loss of MEN1 on developmental regulators (NEUROG3, ASCL1, PAX4, PAX6, ISL1, NKX2.2, INSM1, ARX, Notch, Wnt, BMP, Shh, MAPK/ERK, mTOR) of duodenal enteroendocrine cells, results in gastrinoma development. She contributed towards all the investigated areas.

Rahimeen Rajpar, MD is a dedicated medical professional with a passion for unraveling the mysteries of disease origins and progression. Currently pursuing her residency in internal medicine. RR is committed to advancing the field of medical research. Her goal is to become a leader in the field of Medicine, looking at the medical intricacies from a different lens. Apart from research RR remains dedicated to improving the lives of her patients through comprehensive care and by working towards scientific discoveries.

**11. Manwar Madhwani (MM)** is the co-author of the study. He contributed to the results and conclusions of the study, also contributed to the writing and editing of these sections. He also contributed to the references. He contributed to investigating how the loss of MEN1 on developmental regulators (NEUROG3, ASCL1, PAX4, PAX6, ISL1, NKX2.2, INSM1, ARX, Notch, Wnt, BMP, Shh, MAPK/ERK, mTOR) of duodenal enteroendocrine cells, results in gastrinoma development. He contributed towards all the investigated areas.

Manwar Madhwani, MBBS - Sindh Medical College – Jinnah Sindh Medical University, Karachi, Pakistan. He is currently a medical officer in an NGO. He is interested in research in disease development mechanisms. His goal is to do Medical Residency in Internal Medicine after completing his ECFMG Certification. He aspires to be a specialist in Cardiology/ Pediatrics.

**12. Faryal Yaqoob (FY)** is the co-author of the study. She contributed to the results and conclusions of the study, also contributed to the writing and editing of these sections. She also contributed to the references. She contributed to investigating how the loss of MEN1 on developmental regulators (NEUROG3, ASCL1, PAX4, PAX6, ISL1, NKX2.2, INSM1, ARX, Notch, Wnt, BMP, Shh, MAPK/ERK, mTOR) of duodenal enteroendocrine cells, results in gastrinoma development. She contributed towards all the investigated areas.

Faryal Yaqoob, Doctor of Pharmacy - Faculty of Pharmacy and Pharmaceutical Sciences, Ziauddin University, Karachi Pakistan. Her goal is also to translate emerging findings in research of disease development mechanisms/ origins into their therapeutic implications.

*The work and contributions of everyone have been described in detail, the order is randomized and the numbering is just for referencing purpose.*

## Figures

**Fig 1.**
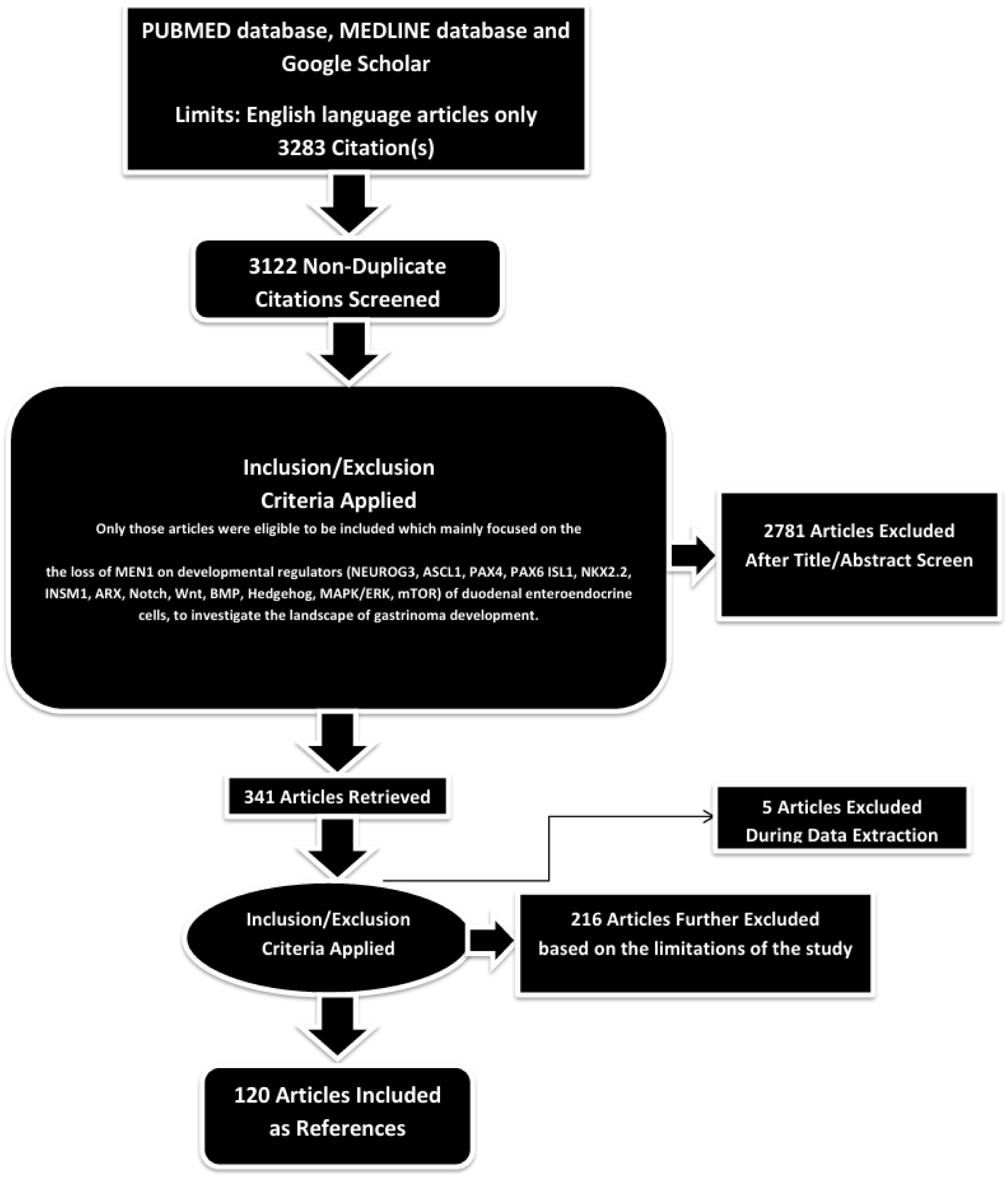
PRISMA FLOW DIAGRAM: This figure represents graphically the flow of citations in the study.

## Notes

### Competing Interest Statement

The authors have declared no competing interest.

